# Which factors had the greatest impact on the United States COVID-19 outbreak? An ecological assessment of mitigation behavior and policy contributions to reducing SARS-CoV-2 transmission in the US from September 2020 through November 2021

**DOI:** 10.1101/2023.07.19.23292882

**Authors:** Velma K Lopez, Sarah Kada, Pragati V Prasad, Taylor Chin, Betsy L. Cadwell, Jessica M Healy, Rachel B Slayton, Matthew Biggerstaff, Michael A Johansson

**Author notes:** Corresponding author: Velma K. Lopez.

## Abstract

*Introduction*: United States’ jurisdictions implemented varied policies to slow SARS-CoV-2 transmission. Understanding patterns of these policies alongside individual’s behaviors can inform effective outbreak response.

*Methods*: We estimated the time-varying reproduction number (R_t_), a weekly measure of real-time transmission using US COVID-19 reported cases from September 2020-November 2021. Using two multi- level regression models, we then assessed the association between R_t_ and policies, personal COVID-19 mitigation behaviors, variants, immunity, and social vulnerability indicators. First, we fit a model with state-level policy stringency according to the Oxford Stringency Index (OSI), a composite indicator reflecting the strictness of COVID-19 policies and strength of pandemic-related communication. Our second model included a subset of specific policies.

*Results*: Implementation of stringent observed policies (defined by OSI) was associated with an R_t_ reduction of 6.7- 11.6% (95% Credible Interval [CI]). In the Individual Policy Model, mask mandates had a null association with R_t_ (95% CI: -1.5-0.2%), while other policies were associated with modest reductions: cancellation of public events 95% CI: 1.4-3.7%; restrictions on gathering sizes 95% CI: 0.1- 2.2%; stay-at-home orders 95% CI: 0.3-4.8%. The association between R_t_ and other covariates was similar in both models. Among personal COVID-19 mitigation behaviors in the OSI Model, R_t_ was estimated to decrease 18%-26% (95% CI) with a 50% reduction in national airline travel, 2.4-3.3% (95% CI) with a 10% reduction in local movement to recreation and retail locations, and 12-15% (95% CI) with self-reported mask use of 50%. Increased COVID-19 seroprevalence and vaccination were both associated with reduced R_t_, 28-32% (95% CI) and 20-23% (95% CI) reductions, if half of people had been previously infected or fully vaccinated, respectively in the OSI Model.

*Conclusion*: SARS-CoV-2 transmission was reduced by layered measures. These results underscore the need for policy, behavior change, and risk communication integration to reduce virus transmission during epidemics.

**Key messages:** <u>What is already known on this topic:</u> When the United States responded to the COVID-19 outbreak, jurisdictions took varied approaches to balance economic well-being with health and safety. Yet, the impact of the pandemic was large - millions of people were infected and over a million people died – and the relative roles of policies, policy-independent behavior change, and other factors remains unclear.

What this study adds: A retrospective analysis of policy implementation and SARS-CoV-2 transmission dynamics over a year and a half indicated that social behavioral change was critical for limiting transmission prior to increases in immunity due to infection and vaccination.

How this study might affect research, practice or policy: While policies contributed to slowing the spread of COVID-19, the largest impact on transmission early in the pandemic was due to individual behavior change, highlighting the importance of identifying and communicating effective control practices regardless of specific policies.

## Main Text Introduction

As SARS-CoV-2 began to spread in early 2020, there was limited knowledge about transmission dynamics and uncertainty regarding the most effective mitigation tools and how to best implement them. Governments began to implement travel restrictions and stay-at-home orders to reduce contact between individuals. These non-pharmaceutical intervention (NPI) policies rapidly evolved and eventually encompassed a diverse range of strategies. NPI policies, such as stay-at-home orders, closure of public facilities, limiting gathering size, and closing schools or universities, and their combinations indeed slowed SARS-CoV-2 spread during the early pandemic phase [1–3]. While NPI policies, particularly those targeting containment of the virus, were markedly successful at reducing transmission opportunities, several exogenous factors also played important roles.

Social structure dramatically affects how people interact and cannot be ignored in disease dynamics. Within the United States (US), physical distancing by staying home from work was a policy option available to more wealthy and White individuals rather than frontline, essential worker populations [4], who are more likely to have a lower socioeconomic status and belong to racial minoritized communities [5,6]. Additionally, SARS-CoV-2 testing sites may have been disproportionately available in areas that had a greater proportion of White residents [7–9]. In other words, the opportunity to know one’s infection status and limit interactions with others, if necessary, was only available to a privileged subset of the population. Moreover, weather added further complexity to these dynamics. For example, SARS-CoV-2 transmission may decrease as temperature and specific humidity increases [10]; potentially weakening the viral envelope [11] and impacting when people spend time indoors, i.e., conditions more favorable to viral transmission. Whether COVID-19 mitigation policies effectively reduced transmission thus requires an assessment within the larger context of behavior, social factors, and weather conditions.

COVID-19 mitigation policies are broadly accepted in the literature as successfully reducing SARS-CoV-2 transmission. However, the relative contribution of policy over long periods of time, while accounting for exogenous factors that also drive transmission, is unknown. Disentangling competing drivers of US SARS-CoV-2 dynamics over time can highlight opportunities to improve future public health outbreak responses. US COVID-19 mitigation policies were diverse and heterogeneously implemented [12].

Additionally, the 55 million cases and over 820,000 deaths reported by December 31, 2021 [13], were spatially patterned by existing demographics and health disparities across jurisdictions. Given these patterns, we seek to understand the relative impact of COVID-19 mitigation policies on US SARS-CoV-2 transmission as both the pandemic and pandemic response evolved.

## Methods

We conducted an ecological assessment of jurisdictional-level SARS-CoV-2 transmission in the US. The analysis period (64 weeks; September 6, 2020, to November 27, 2021) and the fifty-one jurisdictions (all US states and the District of Columbia; DC) analysed were selected for data availability.

*COVID-19 Time-varying Reproduction Number Estimation*: We estimated R_t_, a weekly measure of real-time transmission, using COVID-19 cases reported to the Centers for Disease Control and Prevention (CDC). Onset dates were back-projected from case report dates using time-specific delays; infection dates were sampled using a log-normal distribution for the incubation periods (log mean = 1.63 and log standard deviation = 0.5 based on published data [14]). For each trajectory, we estimated R_t_ using the methods described in Cori et al. [15]. We used a 7-day window and an uncertain serial interval (SI) (mean: 5 days, standard deviation: 1 day), with 5 samples from the SI distribution and 5 samples of the R_t_ posterior for each SI value [16]. We generated 250 R_t_ samples for each time point and jurisdiction and used the mean estimate on each Wednesday for this analysis.

*Covariate data:* We included data on mitigation policies [17], personal mitigation behaviors [18,19], key variants circulation [20], weather [21,22], immunity indicators [23,24], and vulnerability indicators [25]. Standardized policy data were obtained from the Oxford COVID-19 Government Response Tracker [17]. The dataset includes a composite indicator, the Oxford Stringency Index (OSI), which reflects the overall strictness of COVID-19 policies and strength of pandemic-related communication, as well as individual policies. All data are described in the Supplemental Text. Descriptive statistics to inform model building are shown in Supplemental Figures 1-5. We assessed correlation between covariates by estimating the median pairwise R^2^ with bivariate regression models (see Supplemental Figure 1).

*Analytical Approach*: We assessed the association between R_t_ and selected determinants with Bayesian Gaussian multi-level regression models, using a log-link function and jurisdiction- and time-specific intercepts. We assessed two models. The first focused on general government response, using the OSI, a composite indicator of several different policies and communication strategies. Because we were also interested in the individual impacts of some of the specific policies, we built a second model that omitted the general OSI variable and included only a few specific policies that are components of OSI. The second model included four individual policies: cancelation of public events, restrictions on gathering sizes, stay-at-home orders, and mask mandates. Both models were adjusted for the aforementioned covariates, which were selected for inclusion because of their *a priori* relationships with COVID-19 transmission (see Supplemental Text for model statements, samples run, and information about priors). Model convergence was assessed using the Gelman-Rubin convergence diagnostic 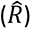 and model fit was evaluated from the predictive posteriors. Jurisdiction-specific results for the OSI model are presented in Supplemental Figure 6.

We conducted several sensitivity analyses. First, we compared these models to the same two models with naïve priors via leave-one-out (LOO) cross-validation [26] (Supplemental Figures 7) and compared models with different structures for temporal correlation (Supplemental Figure 8). Second, we re-ran the primary models using publicly available R_t_ estimates from the Centre for Mathematical Modeling of Infectious Diseases COVID modelling group [27] and compared the model results to those presented here (Supplemental Figure 9). Third, we re-ran the individual policy model without the behavior covariates (Supplemental Figure 10).

Analyses were conducted using R (version 4.2.1), with the *rstanarm* package used for primary analyses. R code is available in a public repository (https://github.com/cdcepi/COVID-19-Mitigation_Rt).

This activity was reviewed by CDC to be deemed not human subject research and therefore a human subject review was not required. The study was conducted consistent with applicable federal law and CDC policy§.

## Results

R_t_ estimates exhibited spatiotemporal variability between September 2020 and November 2021 (Figure 1). All jurisdictions experienced sustained transmission increases (R_t_ > 1) in late 2020, followed by a period of fluctuations until the summer 2021 Delta wave when many had their highest R_t_ estimates. R_t_ dynamics differed substantially between jurisdictions, such as transmission timing and rate increases in late 2020 during the Alpha wave. The lowest mean estimated R_t_ over this period was for Vermont in May 2021 (R_t_=0.66) and the highest value was for DC in November 2021 (R_t_=1.62).

**Figure 1.**
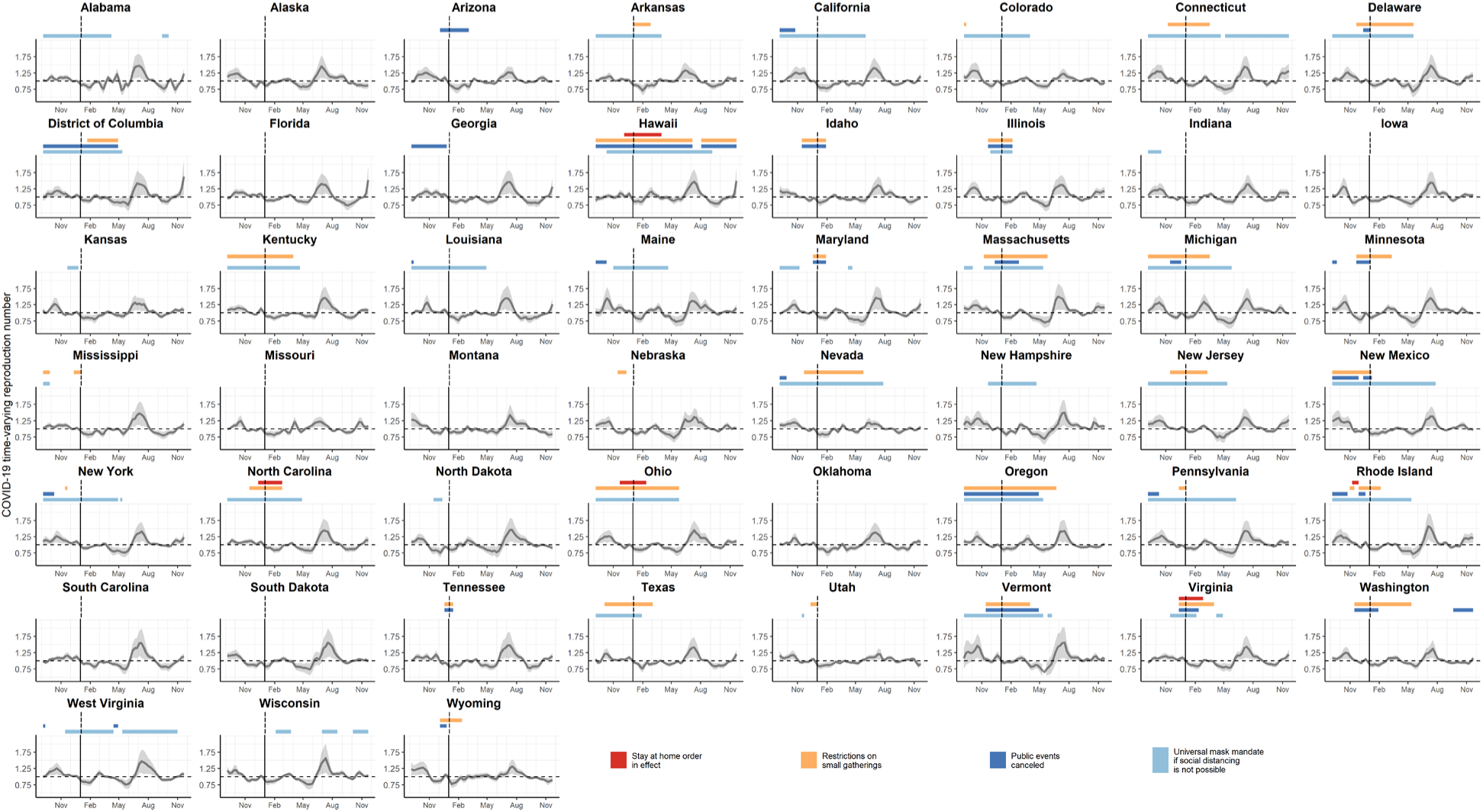
The jurisdiction-specific weekly COVID-19 time-varying reproduction number (R_t_), 90% confidence intervals in grey, and time periods reflecting implementation of specific non-pharmaceutical interventions (NPIs). State-wide implementation of stay-at-home orders are in red, restrictions on gatherings in yellow, cancellation of public events in dark blue, and universal masking when physical distancing was not possible in light blue. The solid vertical line represents January 1, 2021 and the dashed horizontal line reflects an R_t_ value of 1.0.

All individual policies exhibited spatiotemporal variability (Figure 1) and moderate correlation (maximum: 0.47, Supplemental Figure 1B). Overall, stringency of mitigation policies dipped slightly in October 2020 and decreased substantially between March and June 2021 (Figure 2A). Jurisdictional variation in policy stringency persisted, with the lowest median value in South Dakota (0.09; range: 0.06- 0.21) and the highest median value in Hawaii (0.66; range: 0.44-0.76).

**Figure 2.**
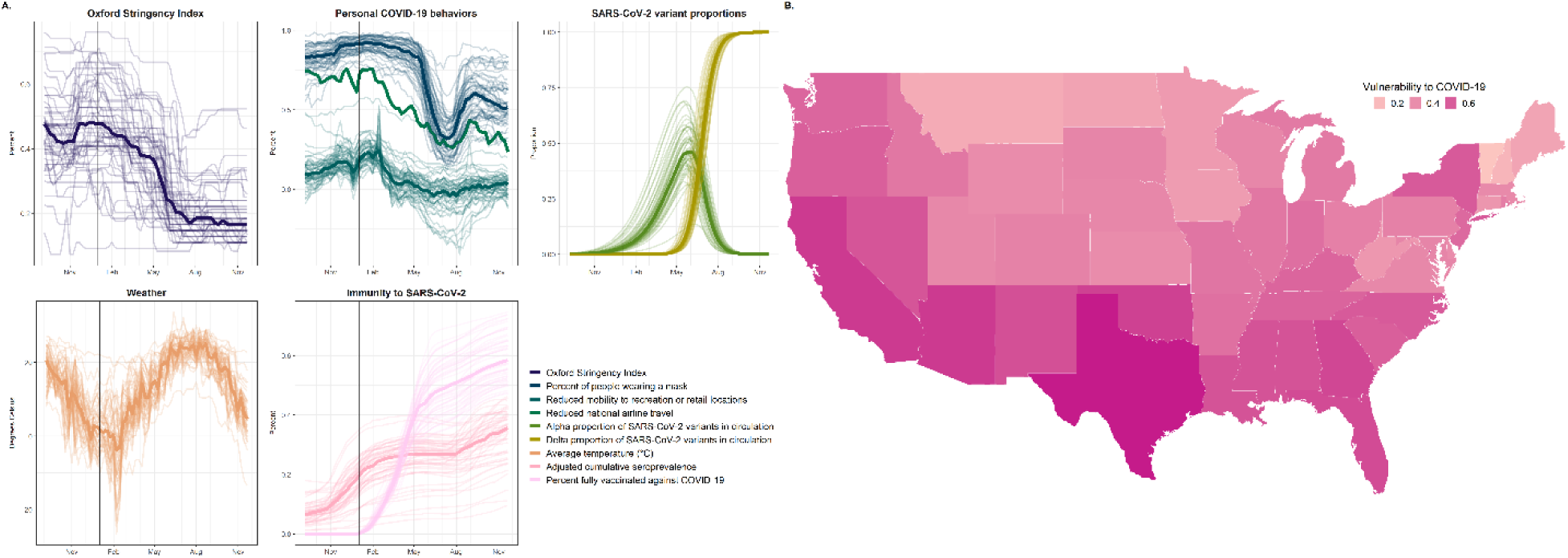
**A**. Distribution of time-varying covariates, including the Oxford Stringency Index (OSI), personal COVID-19 behaviors, proportion of key SARS-CoV-2 variants in circulation, weather, and immunity to SARS-CoV-2. Lines reflect jurisdiction-level observations over time, with the median values across jurisdictions depicted by the bold line. The solid, black vertical line represents January 1, 2021. **B**. Average of the three selected Community Covid-19 Vulnerability Index (CCVI) indicators for each jurisdiction.

Personal COVID-19 mitigation behaviors also varied over time and space (Figure 2A). Reductions in national airline travel and individual mobility showed similar patterns over time, though individual mobility also varied by jurisdiction. Reductions in both were substantial and relatively static until March 2021. In August 2021, all three behavior indicators increased, with new reductions in national airline travel and local mobility, as well as increased reported mask use (Figure 2A). The increased personal mitigation behaviors in August 2021 coincided with the rapid increase in prevalence of the Delta variant (Figure 2A, Supplemental Figure 1A). Expansion of the Alpha variant in early 2021 was generally slower, more heterogeneous, and not correlated with increased mitigation behavior.

There was high heterogeneity in jurisdictional SARS-CoV-2 seroprevalence. By the end of November 2022, seroprevalence ranged from 0.10 in Vermont to 0.46 in Wyoming (Figure 2A). The proportion of the population that was fully vaccinated with an initial vaccine series increased across all jurisdictions beginning with the vaccination distribution in early 2021 and ranged from 0.47 (Alabama) to 0.75 (Vermont) at the end of November 2021.

The average of the five Community COVID-19 Vulnerability Index (CCVI) indicators showed higher vulnerability in southern jurisdictions (Figure 2B).

We fitted two regression models to assess the relationship of these factors with R_t_ dynamics: the OSI Model and the Individual Policy Model. For both models, we assessed alternative spatiotemporal model structures and found that a model with independent random effects for time and state provided the best fit (see Supplement 8). With moderate values for CCVI indicators (0.5 each) and temperature (12°C), estimated R_t_ in the absence of mitigation (i.e., the fixed effect intercept) was 2.2 for the OSI Model (95% Credible Interval [CI]: 2.0-2.5) and 2.5 (95% CI 1.9-3.4) for the policy model.

Overall stringency, some of the specific individual policies, each behavioral component, and both immunity indicators were associated with decreased R_t_ (Figure 3). Implementation of the strictest policies observed in the US relative to least strict policy implementation decreased R_t_ by 9.2% (95% CI: 6.7- 11.6%). Within the individual policies model, cancellation of public events decreased R_t_ by 2.6% (95% CI: 1.4-3.7%), restrictions on gathering sizes by 1.2% (95% CI: 0.1-2.2%), and stay-at-home orders by 2.6% (95% CI: 0.3-4.8%). Mask mandates had a mean estimate corresponding to a 0.7% reduction in R_t_ but did not reach statistical significance (95% CI: -1.5-0.2%). Strong associations were also observed for personal mitigation behaviors in both models. For the OSI Model, R_t_ decreased by 22% (95% CI: 18%- 26%) if there were a 50% reduction in national airline travel, 2.9% (95% CI: 2.4-3.3%) if local movement to recreation and retail locations decreased by 10%, and 14% (95% CI: 12-15%) if self-reported mask use reached 50%. Seroprevalence and vaccination were both associated with reduced R_t_ - a 30% (95% CI: 28- 32%) and 22% (95% CI: 20-23%) estimated reduction if half the population had been previously infected or fully vaccinated, respectively.

**Figure 3.**
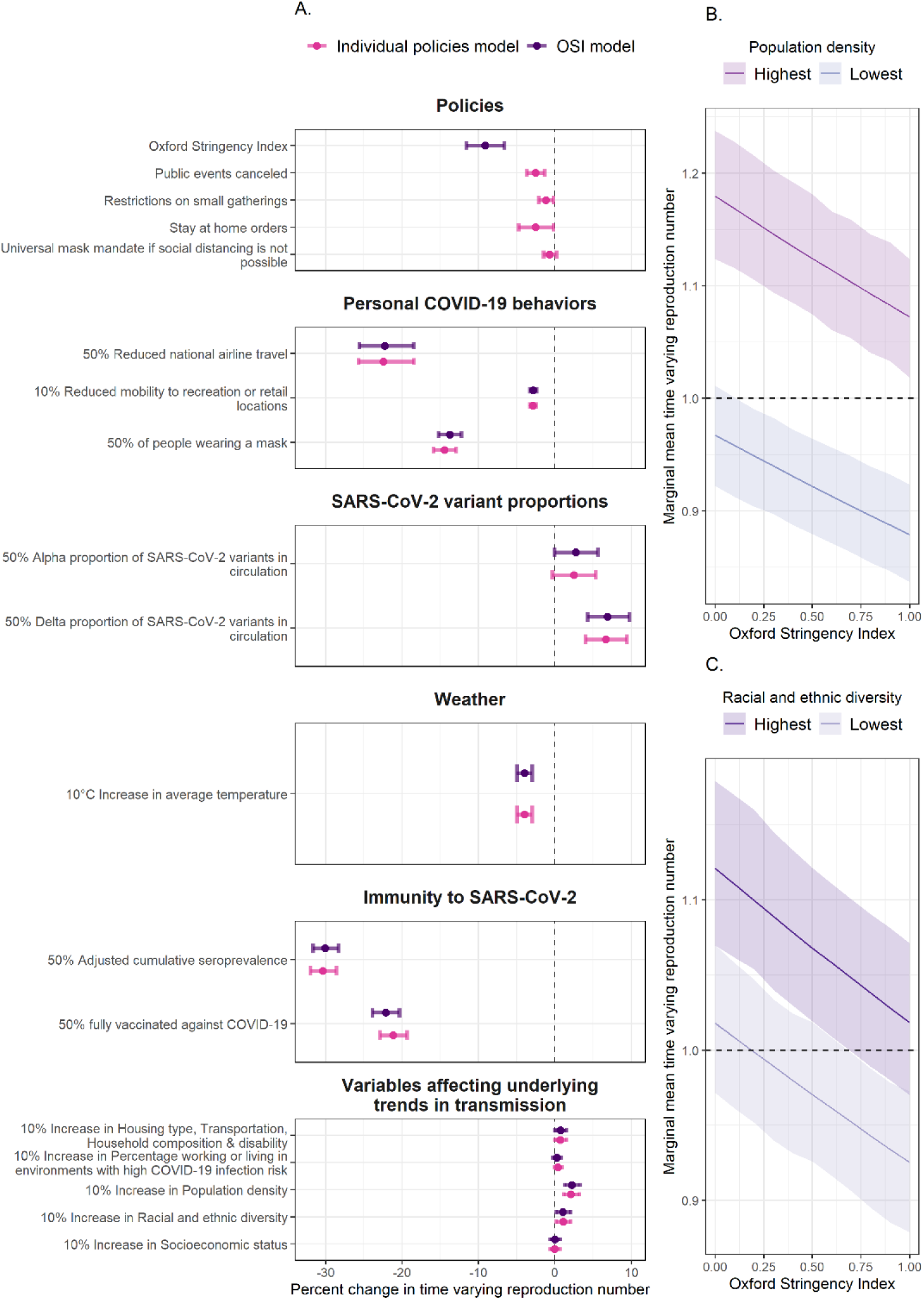
**A**. Percent change in the COVID-19 time-varying reproduction (R_t_) number from policies, personal COVID-19 behaviors, proportion of key SARS-CoV-2 variants in circulation, weather, immunity to SARS- CoV-2, and variables affecting underlying trends in transmission. The results from two different linear regression models are shown below. Model one, in pink, included the Oxford Stringency Index (OSI). Model two, in purple, included a subset of policies used to comprise the OSI. Both regression models were gaussian with log link function and had jurisdiction and time specific intercepts. The posterior mean is represented by the point and the 95% credible intervals by the bars. **B**. The marginal mean in the COVID- 19 R_t_ across the range of observed OSI values in jurisdictions with the highest and lowest population density. **C**. The marginal mean COVID-19 R_t_, and 95% credible interval show in bands, across the range of observed OSI values in jurisdictions with the highest and lowest racial and ethnic diversity.

Community vulnerability also played an important role in transmission. Both population density and a community’s racial and ethnicity diversity level were associated with increases in R_t_ (Figure 3). However, the level of policy stringency needed to reduce R_t_ to less than 1 differed within each of these groups. For example, in jurisdictions with the highest population density (Figure 3B) and the most racial and ethnic diversity (Figure 3C), even the highest OSI values were not sufficient to reduce transmission.

Sensitivity analysis showed consistent estimates for policy impacts even when personal mitigation behaviors were excluded (Supplement 10). Coefficients were similar in direction for both models, with or without informative priors (see Supplement 6), and minor magnitude differences were observed in our sensitivity analyses with alternative R_t_ estimates (Supplement 9).

We estimated the proportional reduction associated with each time-varying component individually and combined for each jurisdiction and nationally over time using the OSI Model estimates (Figure 4, Figure 5, and Supplemental Figure 6). Over the study period, personal mitigation behaviors were associated with the largest proportional transmission reductions in all jurisdictions before vaccine implementation and remained an important or leading contributor to reduction thereafter (median combined reduction over time across locations: 44%, range: 10-62%) (Figure 5). Immunity was the second most important contributor overall and of growing importance as more people were infected and vaccination coverage increased. These patterns, however, were starkly different between jurisdictions (Figure 4 and Supplemental Figure 6). For example, in November 2021 many jurisdictions had higher estimated reductions associated with previous infections than vaccination (e.g., Wyoming). Meanwhile, other jurisdictions with lower seropositivity or more vaccinations had higher estimated reductions associated with vaccination (e.g., Vermont). At the end of the study period, SARS-CoV-2 seroprevalence and vaccination were associated with a wide range of jurisdictional reductions (8-35% and 24-36%, respectively). Policies and weather were also associated with transmission changes but with less overall estimated magnitude than the effects of behavior and immunity.

**Figure 4.**
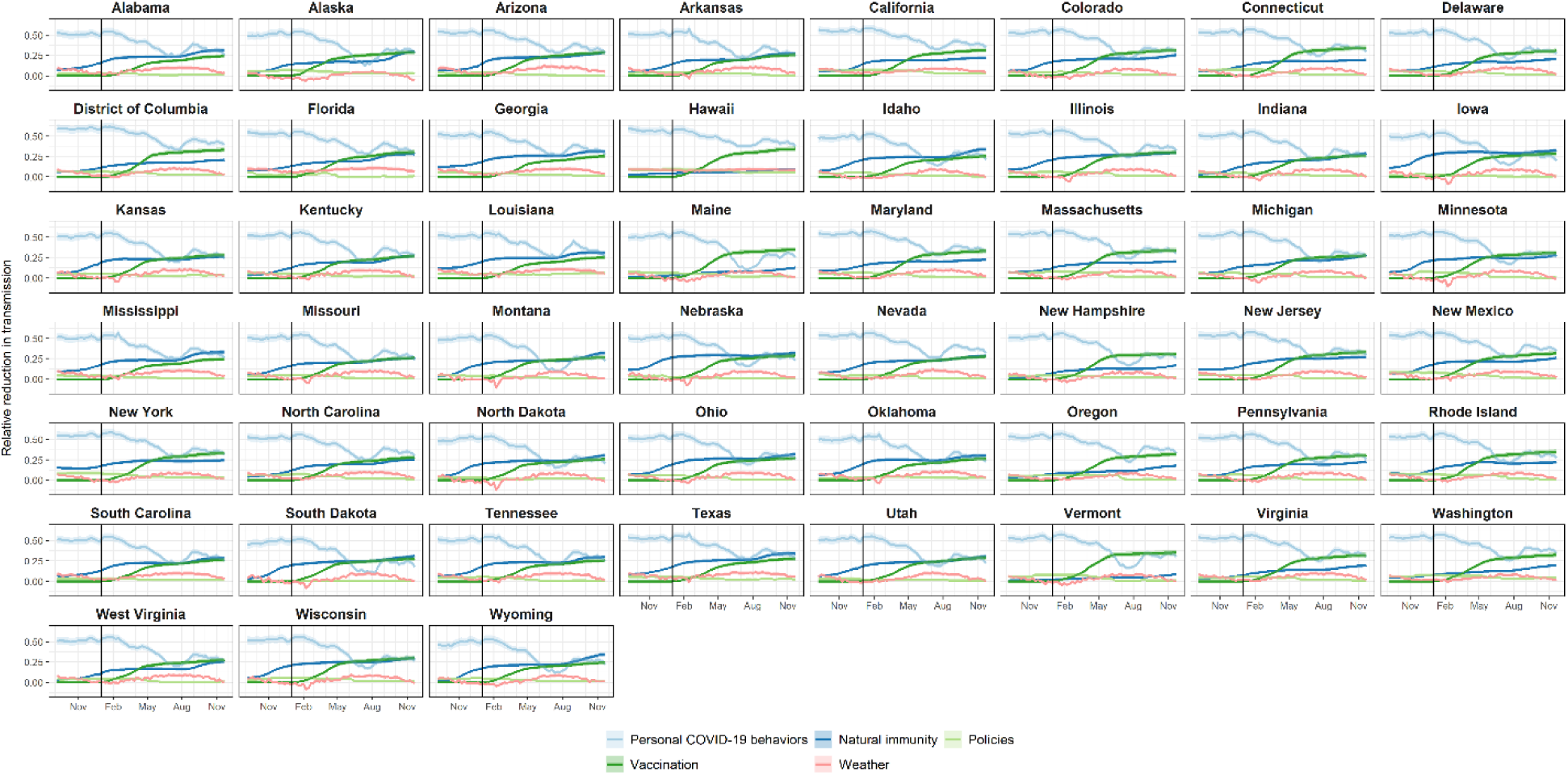
Estimates for jurisdiction-level proportional reductions in the COVID-19 time-varying reproduction number (R_t_) over time for the Oxford Stringency Index (OSI) regression model. Colored lines depict the relative reduction for select sets of covariates, with the 95% credible interval in the corresponding-colored bands, and the solid vertical line represents January 1, 2021.

**Figure 5.**
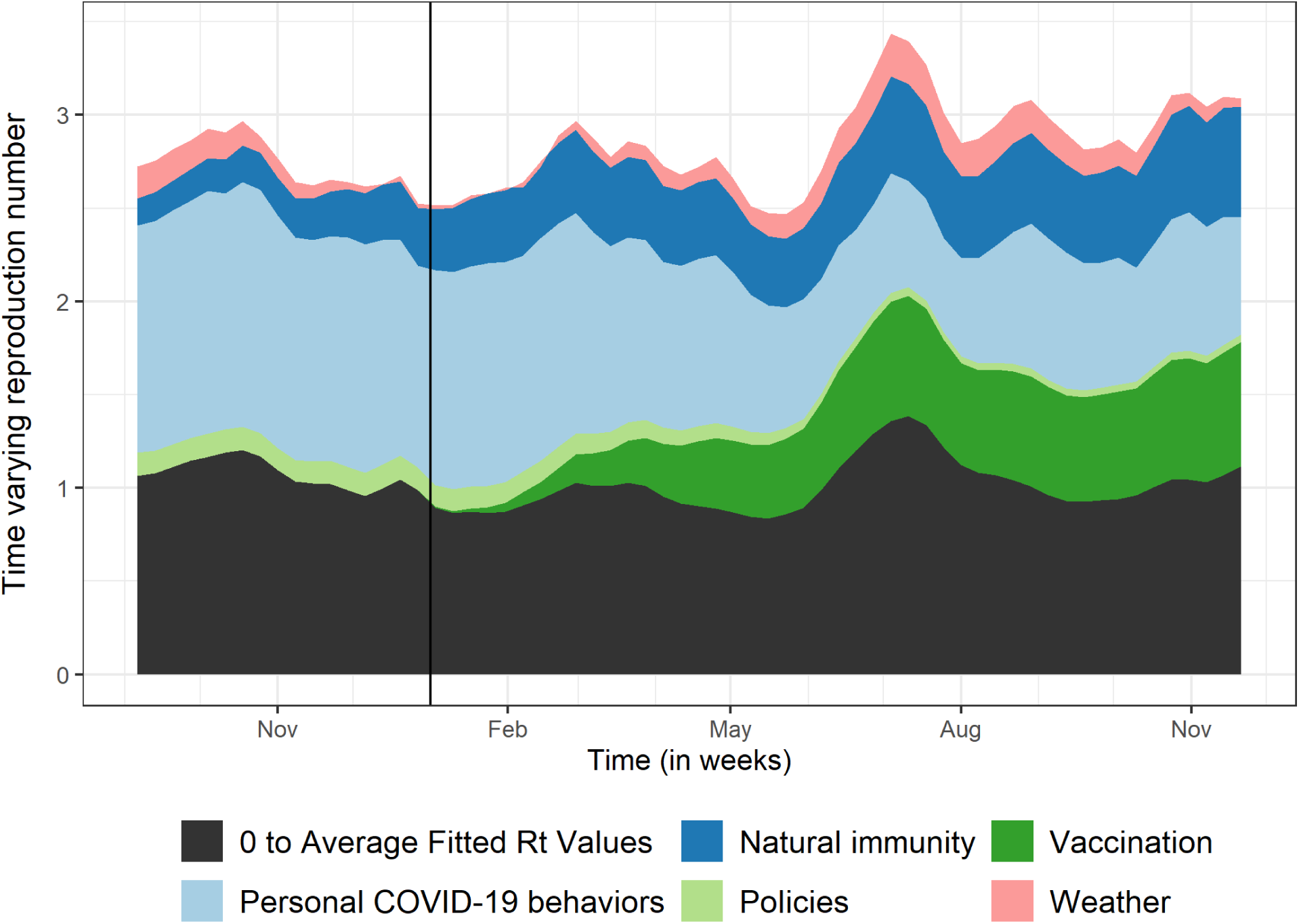
Average relative contribution of observed covariates on fitted COVID-19 time-varying reproduction number (R_t_) across all jurisdictions from the OSI model. The highest values over time (i.e., the top of the pink band) represent R_t_ estimates including only the effects of local vulnerability and variants. The pink band represents reductions in R_t_ associated with changing weather. The blue bands represent reductions in R_t_ associated with infection-acquired immunity (dark blue) and with behavior modification (i.e., masking use, mobility, and national airline travel, combined and depicted with light blue). The green bands represent reductions in R_t_ associated with policies (light green) and vaccination (dark green). The fitted values from the regression model are represented with the top of the black band. The solid vertical line represents January 1, 2021.

## Discussion

Deciphering SARS-CoV-2 transmission drivers throughout the pandemic can inform future mitigation policies and interventions for respiratory pathogens. Our analysis integrates spatial and temporal patterns of potential transmission determinants to assess associations between those determinants and SARS-CoV-2 transmission. We included a wide range of determinants with the goal of assessing the relative impact of each as the pandemic progressed. On aggregate, while both general and specific policies and behavior were associated with reduced SARS-CoV-2 transmission, we found that vaccination uptake, mask wearing, and reduced airline travel were associated with greater transmission reductions than government policies or mandates. Yet, the proportional reduction on transmission varied overtime; mask wearing and reduced airline travel had large impacts early on, while the relative contribution of immunity - due to previous infection and vaccination - increased as the pandemic continued, and behavior modification decreased. These findings imply that behavioral mitigation was critical to limiting transmission, reinforce the value of vaccines in returning to more normal behavior, and indicate the complexity of relationships between infection dynamics, behavioral choices, and government policies.

Early US COVID-19 mitigation strategies focused on physical distancing and masking policies. Our estimates suggest that physical distancing policies reduced SARS-CoV-2 transmission, with an estimated mean reduction of 1-3% for individual policies and an estimated total reduction of approximately 9% if sets of policies tracked by the OSI were at the highest observed levels in the US. These reductions are comparable with other, short-term, national-level assessments of NPI policies and showed reductions in COVID-19 cases [28–31], transmission [32–34], and deaths [29]; there was some reduction in effectiveness for policies with longer durations [29]. In contrast to our null results for masking policies, jurisdiction-specific studies show short-term effectiveness of mask mandates in reducing cases [35] and hospitalization growth rates [36]. Importantly, most of these early evaluations focused on the period in which the NPIs were in effect and did not assess long-term effects or periods when the NPI was not implemented, as was done here. There is some evidence of limited prolonged reduction in COVID-19 outcomes when NPI policies were lifted [37], with more socially disadvantaged communities experiencing greater rebounds in COVID-19 burden than other communities [38]. Overall, the association between individual policies and transmission reductions was strong but accounted for only a modest overall transmission risk reduction. This finding highlights the need to better understand adherence to the policies and what drivers personal behavior mitigations.

The relationships underlying policy implementation and behavior are intricate. Like others, we found indicators of behavior such as local mobility, national airline travel, and self-reported mask use to be associated with significant reductions in R_t_ although not necessarily temporally aligned with the corresponding policies [39]. For example, analysis of early mobility data showed that movement patterns changed drastically even before most physical distancing policies were implemented [40], with continued reductions in movement after policies were in place [41]. It is plausible that broad agreement within the physical distancing policies early in the pandemic [42] influenced personal choices to stay home. Similarly, self-reported mask use increased, and local mobility decreased rapidly as the Delta wave grew even though updated recommendations stated that vaccinated individuals could resume pre-pandemic activities without wearing a mask, once again, indicating a behavioral response that was independent of policy. Conversely, one example of temporal alignment between policy and behavior was immediately following the May 2021 guidance update when there was a rapid decrease in reported mask use and increases in local mobility, likely an effect of individuals returning to activities that had previously stopped due to the pandemic.

Before the Delta wave, US vaccination rates were steadily rising, with varying geospatial, socioeconomic, and race/ethnic coverage patterns [43,44]. Many jurisdictions also experienced substantial transmission in 2020 and early 2021, resulting in greater infection-acquired immunity. While we did not adjust for waning immunity [45] or for changing vaccine effectiveness with the appearance of new variants [45,46], we found associations between infection-acquired immunity and vaccination with decreased transmission. It is plausible that if we adjusted our model estimates for waning immunity, the strength of the immunity-related associations would be attenuated. In most states, the relative impact of either infection-acquired immunity or vaccination was as high as the decreasing impact of behavior change by November 2021, when the impact of vaccination was higher than the impact of infection-acquired immunity. There were also distinct jurisdictional differences, with some jurisdictions showing comparable impact by late 2021 (e.g., Alabama, Arkansas) and others showing much higher impact from vaccination than infection-acquired immunity (e.g., Connecticut, Hawaii). Additionally, we did not adjust our model for COVID-19 testing, which may have a non-linear relationship with seropositivity since testing availability and behaviors changed over time.

Social structure drives transmission patterns for all pathogens and the US COVID-19 epidemic highlighted existing social fault lines that influenced not only who in society was more likely to get infected, but also who was more likely to benefit from mitigation measures [47,48]. Here, we accounted for multiple social vulnerability indicators and found higher transmission rates in states with higher population density and greater racial and ethnic diversity. Stronger mitigation was needed to reduce R_t_ below the critical threshold of 1 for communities with higher vulnerability. This suggest that a one-size- fits all approach to mitigation is not appropriate and that more vulnerable communities may benefit from intentional support. Our findings align with several results from county-level analyses. For example, higher SARS-CoV-2 transmission rates occurred in low-income and racial minoritized communities [49]. Additionally, counties with higher social vulnerability were more likely to become a COVID-19 burden hotspot [50], that is a geographic area with elevated disease incidence [51]. Because structural racism is part of the intersectional factors comprising social vulnerability, it is unsurprising that hotspots were common in US counties with a greater percentage of non-White residents [52–54]. These findings highlight the importance of incorporating social markers of risk in infectious disease transmission models [55].

Our findings do not reflect causal relationships. Establishing causality between mitigation measures and transmission is complicated by a variety of risk factors, from SARS-CoV-2 variants emergence and changes in human behavior to environmental conditions– which all fluctuate over time and space. First, many important SARS-CoV-2 transmission determinants are correlated. To partially address this limitation, we removed highly correlated variables that measured similar factors when possible.

However, we did choose to retain some highly correlated variables, which may have influenced the findings. Determining the impact of these highly correlated variables, however, is not straightforward given the directionality of the conditional associations and the correlations between the covariates themselves. Second, jurisdiction-level policies may differ substantially from policies implemented at a county or city level. Additionally, many different policy variations were implemented (e.g., some jurisdictions required masks universally, whereas others only in certain locations or where physical distancing was not possible), even within jurisdictions, and those variations were not captured here. Third, we applied a regression framework, which assumes log-linear independence between covariates that does not account for the observed correlation between variables. Overall, the potential causal pathways between the predictors and R_t_ are not individually identifiable at this scale.

Ideally, the wealth of data and intervention diversity in the United States could be used to develop specific recipes for control. However, the diversity and correlation between many contributing factors make precise intervention estimates and/or combinations of interventions infeasible. Here, we conducted an ecological analysis of key strategy types and found that personal mitigation behaviors were more strongly associated with decreased transmission than policies. While most policies may not be sufficient to control COVID-19 on their own, a combination of policies and communication efforts that promote, support, and reinforce behavior change may be an essential pathway for mitigation. The other most impactful intervention was vaccination, a nationwide intervention that was not available early on but became as important as behavior modification for controlling transmission in most jurisdictions by mid to late 2021 [29]. Importantly, transmission was reduced not by a single measure, but by various layered measures; this indicates that no single measure is likely to control SARS-CoV-2 on its own [56]. These findings demonstrate the complexity of the COVID-19 response and SARS-CoV-2 transmission and illustrate the ongoing importance of layered mitigation approaches integrated across public health, government, and communities.

## Data Availability

COVID-19 case data are available upon request at https://data.cdc.gov/Case-Surveillance/COVID-19-Case-Surveillance-Restricted-Access-Detai/mbd7-r32t. All other data are in the public domain and referenced in Supplement 1.

## CDC disclaimer

The findings and conclusions in this report are those of the authors and do not necessarily represent the official position of the U.S. Centers for Disease Control and Prevention.

## Data Availability

All data produced in the present work are contained in the manuscript

## Acknowledgments

This research is based on survey results from Carnegie Mellon University’s Delphi Group. Thank you to Prabasaj Paul for his thoughtful comments and review of this manuscript.

## Author Contributions

MAJ and VKL designed the research question, with assistance from SK and JMH. VKL designed the analysis plan, with components run by VKL, SK, PVP, and TC. VKL wrote the manuscript in consultation with BLC, JMH, RBS, MB, and MAJ. All authors provided critical feedback to help shape the analysis and manuscript. All authors accept full responsibility for the finished work and/or the conduct of the study, had access to the data, and controlled the decision to publish.

## Patient and Public Involvement

Because we used anonymous, public data, we did not include individuals that in the design, conduct, reporting, or dissemination plans of this work.

## Ethics Approval

No Ethics Approval needed.

## Footnotes

§ See e.g., 45 C.F.R. part 46, 21 C.F.R. part 56; 42 U.S.C. §241(d); 5 U.S.C. §552a; 44 U.S.C. §3501 et seq.

