## Supplemental Material for "Which factors had the greatest impact on the United States COVID-19 outbreak? An ecological assessment of mitigation behavior and policy contributions to reducing SARS-CoV-2 transmission in the US from September 2020 through November 2021"

### Supplementary Information Text

#### Covariate Data

We included covariate data on COVID-19 mitigation policies, personal COVID-19 mitigation behaviors, the circulation of key SARS-CoV-2 variants, weather data, indicators of immunity (acquired through infection or vaccination), and COVID-19 vulnerability indicators. As described in the Introduction, each of these sets of variables can either directly or indirectly affect community transmission of SARS-CoV-2. Data were publicly available and management procedures were implemented for each of the covariate data types:

COVID-19 mitigation policies: Standardized policy data were obtained from the Oxford COVID-19 Government Response Tracker at https://github.com/OxCGRT/covid-policy-tracker/tree/master/data.

*The Oxford Stringency Index (OSI):* OSI which ranges from 0 (least stringent) to 100 (most stringent), is a weighted average of nine measures over time (school closures, workplace closure, cancellation of public events, restrictions on gatherings, closed public transport, stay-at-home orders, restrictions on internal movement, international travel control, and public information campaigns). Weights were used to account for whether specific policies were implemented across an entire jurisdiction or only within geographical subunits. We used the smooth version of the OSI variable, which interpolates missing values, within the analysis. We rescaled the values to range from 0 to 1 and calculated a jurisdictional weekly mean from the daily timeseries. For the regression models, we normalized the variable so that the largest observed value was equal to 1.0.

*The four policy variables included in the OSI dataset*: We chose to examine these four policies because they were commonly implemented across the United States and represented key, distinct mitigation measures: cancellation of public events, restrictions on gathering sizes, stay-at-home orders, and mask mandates. We dichotomized all policy variables into the strictest policy versus all other implementations/no policy.

In the original dataset, each of the 4 policy variables were ordinal and on a daily time step and included a second associated variable that described the geographic unit of implementation. We recoded each of these variables following two steps. First, if a policy was implemented in a subunit and not across the whole jurisdiction, we coded the original policy as ‘no jurisdiction-level policy’ (0). Second, we examined the times series for each of the four policies. We converted daily data to weekly data by setting the weekly value to the most common daily value for each week, defaulting to a stricter value in the case of ties if more than two policies were recorded in a given week. Next, we dichotomized all three OSI policy variables so that the strictest level was coded as 1 [strictest policy in effect] and all other levels coded as 0 [weak or no jurisdiction-level policy in effect]. For example, using this schema, the following policy implementations were coded as 1: cancellation of all public events, restrictions on gatherings of 10 people or less, and stay-at-home orders that only allowed leaving the house for essential activities. We coded mask mandates as 1 if masking was required in all public spaces or required in all public spaces where physical distancing was not possible; all other categories of mask mandates (masking required in some areas if physical distancing was not possible, masking was recommended, or no policies) were coded as 0.

Personal COVID-19 mitigation behaviors: Jurisdiction-level, personal behavior data were collected from a variety of sources. Self-reported mask use in public (previous 5 or 7 days) and attendance at gatherings (in the past 24 hours) were collected from the COVID-19 Trends and Impact Survey of Facebook users (https://cmu-delphi.github.io/delphi-epidata/symptom-survey/;) (See **Supplemental Figure 2**); mobility data were collected from Google's COVID-19 Community Mobility Reports (https://www.google.com/covid19/mobility/); and national travel estimates were collected from the Transportation Security Administration (https://www.tsa.gov/coronavirus/passenger-throughput). From the Community Mobility Report data, we included the proportional reduction in weekly median mobility to retail and recreation locations relative to baseline mobility from January 3- February 6, 2020. We also included the weekly median reduction in national airline travel relative to maximum weekly travel in 2019. We set the reference to the maximum weekly travel in 2019 to ease interpretation of the coefficients in the final model.

Circulation of key SARS-CoV-2 variants: We estimated the weekly proportion of Alpha (B.1.1.7) and Delta (B.1.617.2) SARS-CoV-2 variants in circulation by fitting sequence data to a multinomial logistic regression model, which included normalized survey weights to account for reporting patterns within and between jurisdictions[1].

Weather data: We pulled temperature (°C) data from weather stations included in the National Oceanic and Atmospheric Administration's Integrated Surface Database[2], using the package "worldmet" (https://davidcarslaw.github.io/worldmet/). From station level data, we calculated the weekly median temperature in each jurisdiction. We replaced missing observations with the average value of the week before

[1] Paul P, France AM, Aoki Y, Batra D, Biggerstaff M, Dugan V, et al. Genomic Surveillance for SARS-CoV-2 Variants Circulating in the United States, December 2020–May 2021. MMWR Morb Mortal Wkly Rep. 2021;70(23):846–50

[2] Global Hourly - Integrated Surface Database (ISD) | National Centers for Environmental Information (NCEI) [Internet]. [cited 2022 Dec 2]. Available from: https://www.ncei.noaa.gov/products/land-based-station/integrated-surface-database

and after. Given the role of humidity in respiratory virus transmission [3,4], we also assessed associations with relative humidity and absolute humidity to guide our modeling (**Supplemental Figure 3**).

Infection-acquired immunity: We included infection-acquired and vaccine-derived immunity to SARS-CoV-2 indicators in our models. As a proxy measure of infection-acquired immunity, we modeled jurisdiction-level seroprevalence, adjusting estimates for reduced percent of positive assays based on waning immunity using methods described by García-Carreras and colleagues [5] using data from national SARS-CoV-2 serosurveys (https://data.cdc.gov/Laboratory-Surveillance/Nationwide-Commercial-Laboratory-Seroprevalence-Su/d2tw-32xv). Because data were unavailable for North Dakota and DC, we imputed weekly seroprevalence estimates by averaging values from the jurisdictions surrounding these two locations (Minnesota, Montana, and South Dakota, and Maryland and Virginia, respectively). For vaccination, we used the weekly jurisdictional percentage of individuals with a completed primary series of COVID-19 vaccine https://data.cdc.gov/Vaccinations/COVID-19-Vaccination-Trends-in-the-United-States-N/rh2h-3yt2.

COVID-19 vulnerability indicators: We included variables that represent static underlying components that influence transmission, which were developed as part of the Community COVID-19 Vulnerability Index (CCVI) (https://precisionforcovid.org/ccvi) and range from 0 to 1 across all jurisdictions: 1) Racial and Ethnic Diversity 2) Percentage of Population Working or Living in Environments with High COVID-19 Infection Risk, 3) Socioeconomic Status, 4) Housing type, Transportation, Household Composition and Disability, and 5) Population Density (see **Supplemental Figure 4** for Pearson correlation coefficients for CCVI indicators, and **Supplemental Figure 5** for spatial distribution and correlation with time-varying covariates).

Following data management, we explored correlation between all covariates in the data set (**Supplemental Figure 1**).

**Models and posteriors**

The two primary models in this analysis assess the association between $R_t$ and COVID-19 mitigation policy with Bayesian Gaussian hierarchical regression models, using a log link function and jurisdiction-specific intercepts:

$$\text{Model 1 [the OSI Model]}: log\, log\left(Rt_{\,i,j}\right) = \beta_0 + \beta_1 OSI_{i,j} + \beta_2\, SARSCoV2\, seroprevalence_{i,j} + \beta_3 Racial\ and\ Ethnic\ Diversity_{\ i} + \beta_4 Percentage\ of\ Population\ Working\ or\ Living\ in\ Environments\ with\ High\ COVID-19\ Infection\ Risk_i + \beta_5 Socioeconomic\ Status_i + \beta_6 Housing\ type, Transportation, Household\ Composition\ \&\ Disability_i + \beta_7 Population\ Density_i + \beta_8 Mobility_{i,j} + \beta_9 Percent\ fully\ vaccinated_{i,j} + \beta_{10} Mask\ use_{i,j} +$$

[3] Shaman J, Pitzer V, Viboud C, Lipsitch M, Grenfell B. Absolute Humidity and the Seasonal Onset of Influenza in the Continental US. PLoS Curr [Internet]. 2009 [cited 2022 Dec 2];2(DEC). Available from: /pmc/articles/PMC2803311/

[4] Ma Y, Pei S, Shaman J, Dubrow R, Chen K. Role of meteorological factors in the transmission of SARS-CoV-2 in the United States. Nat Commun 2021 121. 2021 Jun 14;12(1):1–9.

[5] García-Carreras B, T Hitchings MD, Johansson MA, Biggerstaff M, Slayton RB, Healy JM, et al. Accounting for assay performance when estimating the temporal dynamics in SARS-CoV-2 seroprevalence in the U.S. [cited 2022 Dec 2]; Available from: https://doi.org/10.1101/2022.09.13.22279702

$$\beta_{11}Temperature_{i,j} + \beta_{12}National\ airline\ travel_{i,j} + \beta_{13}Proportion\ of\ alpha\ variant\ in\ circulation_{i,j} + \beta_{14}Proportion\ of\ delta\ variant\ in\ circulation_{i,j} + b_i + b_j + \varepsilon_{i,j}$$

Model 2 [the Individual Policy Model]: $log\ log\ (Rt_{\ i,j}) = \beta_0 + \beta_1 Public\ events\ canceled_{i,j} +$

$$\beta_2 Restrictions\ on\ small\ gatherings_{i,j} + \beta_3 Stay\ at\ home\ orders_{i,j} + \beta_4 Universal\ mask\ mandate\ if\ social\ distancing\ is\ not\ possible_{i,j} + \beta_5\ SARSCoV2\ seroprevalence_{i,j} + \beta_6 Racial\ and\ Ethnic\ Diversity_{\ i} + \beta_7 Percentage\ of\ Population\ Working\ or\ Living\ in\ Environments\ with\ High\ COVID-19\ Infection\ Risk_i + \beta_8 Socioeconomic\ Status_i + \beta_9 Housing\ type, Transportation, Household\ Composition\ \&\ Disability_i + \beta_{10} Population\ Density_i + \beta_{11} Mobility_{i,j} + \beta_{12} Percent\ fully\ vaccinated_{i,j} + \beta_{13} Mask\ use_{i,j} + \beta_{14} Temperature_{i,j} + \beta_{15} National\ airline\ travel_{i,j} + \beta_{16} Proportion\ of\ alpha\ variant\ in\ circulation_{i,j} + \beta_{17} Proportion\ of\ delta\ variant\ in\ circulation_{i,j} + b_i + b_j + \varepsilon_{i,j}$$

Where *j* reflects weekly estimates; *i* reflects jurisdiction specific estimates; and *b* reflects independent random effect where $b_i \sim N(0,\sigma^2)$ and $b_j \sim N(0,\sigma^2)$.

As described in the methods, for each model, we ran four Markov chains at 2,500 iterations each, with a burn in period of 1,250 iterations. We specified priors for an expected negative association (OSI, all individual mitigation policies, mobility, masking, reduced airline travel, cumulative COVID-19 cases, vaccination, and temperature) or positive association (variants and each CCVI indicator) between $R_t$ and the covariate of interest. For each coefficient with an expected negative association, we used a normal distribution with a mean of -0.7 and standard deviation of 0.1, approximating a 50% decrease with a 95% interval of 40-60%. We used a normal prior with a mean of 0.4 and a standard deviation of 0.1 for covariates with expected positive associations, approximating a 50% increase with a 95% interval of 20-80%. The intercept prior had a normal distribution with a mean of 1.1 and standard deviation of 0.1, reflecting an expected Rt without any mitigation behaviors or policies from 2.5 to 3.7 (95% interval). Posterior estimates from the regression models are presented in **Supplemental Table 1**. These estimates were used to calculate the percent change in $R_t$ shown in Figure 3.

Sensitivity Analyses

Following the first run of the models, we compared them to the models with naïve priors via leave-one-out cross-validation (LOO) to estimate the estimate expected log posterior predictive density (elpd) and visualize each model's predicted values and residuals (**Supplemental Figure 7** Panel **A** and Panel **B**). We assessed model convergence via $\hat{R}$ values (the Gelman-Rubin convergence diagnostic). The naïve priors used here reflect a specified normal prior with a mean 0, standard deviation of 2.5 for the intercept and all coefficients; while for the standard error, a specified exponential prior with a standard deviation of 1 was used. The scale of the priors was internally adjusted to make them weakly informative.

To determine the most appropriate model structure for the data, we compared three different model structures: a generalized linear model (GLM) with a random intercepts per state, a GLM with random intercepts per state and per week, a GLM with random intercepts per state-week, and a generalized additive model (GAM) with a spline for time and state-specific random intercepts (**Supplemental Figure 8**). Each model included the priors described in the main text. We compared each model's expected log posterior predictive density (elpd) from a 10-fold cross validation (**Supplemental Table 2**), primary trends in time (**Supplemental Figure 8A**), and fixed effect posteriors (**Supplemental Figure 8B**). The GLM with random intercepts for state and time had the highest elpd with no observations indicative of strong influence on the model, suggesting the best fit to the data.

We compared the OSI Model and the Individual Policy Model to a second set of models that used a different data source to estimate $R_t$ (referred to as *epiforecast* $R_t$; see **Supplemental Figure 9**). We pulled all jurisdiction-level $R_t$ files (n=484) from https://github.com/epiforecasts/epiforecasts.github.io [6] and extracted the median $R_t$ estimate for all Wednesdays (mid-week estimate to smooth changes in daily reporting). We then calculated the median $R_t$ per date to generate a weekly estimate (see Panel **A** for $R_t$ estimate comparison) and fit regression models to these data using the same methods described for the primary models (see Panel **B** for posteriors). We compared the models via leave-one-out (LOO) and examined the posteriors, predicted Rt values, and model residuals (Panel **C**).

[6] Abbott S, Hellewell J, Thompson RN, Sherratt K, Gibbs HP, Bosse NI, et al. Estimating the time-varying reproduction number of SARS-CoV-2 using national and subnational case counts. Wellcome Open Res 2020 5112. 2020 Dec 8;5:112.

**Fig S1. A. and B.**

To determine the correlation between each of the unique pairs of variables in the model, we fit bivariate Bayesian multi-level Gaussian regression models, and included a jurisdiction-specific intercepts (as random effects). We exclude variables that did not vary by time or space in this sensitivity analysis; due to the unbalanced nature of the policy data, we also excluded pairs where the policy variables would have been outcome variables in the regression pairs. For each model, we estimated the median $R^2$ values and present the average $R^2$ value for each pair (if applicable) in Panel **A**. In Panel **B**, we present the median $R^2$ values for policy related regression models.

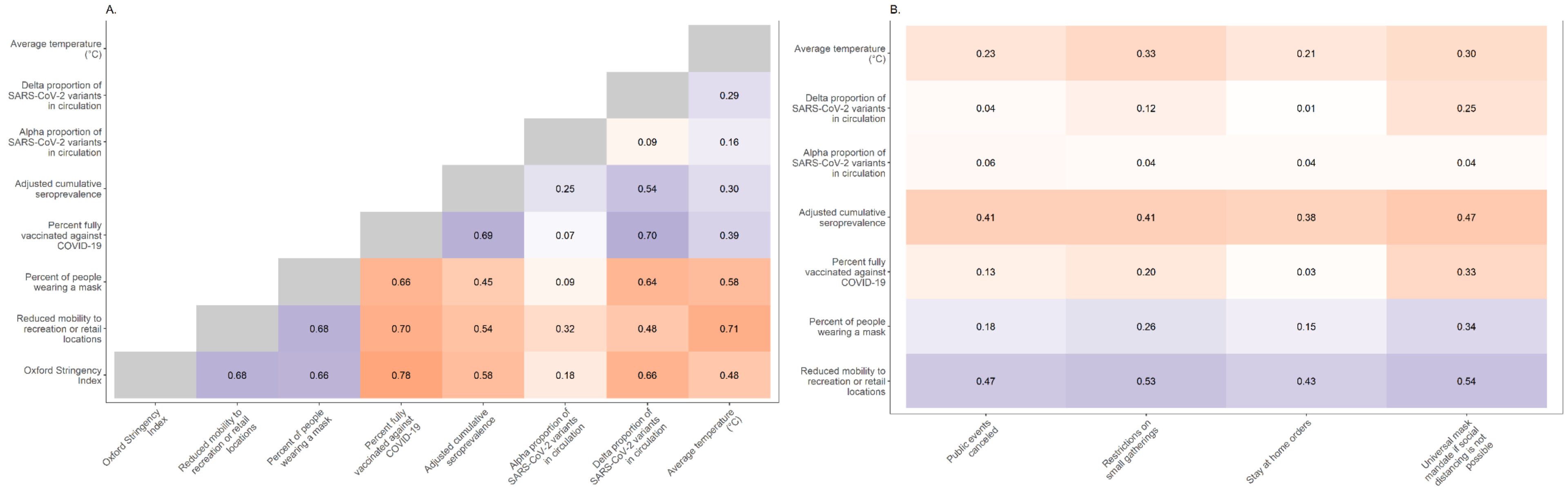

**Fig S2.**

Proportion of Facebook respondents that self-report mask wearing and attending a gathering of 10 or more people. Each line represents one jurisdiction. These variables are highly correlated, with a Pearson correlation coefficient of -0.83 over the entire analysis period and across all jurisdictions. The solid vertical line represents January 1, 2021. Because of the high correlation, we focused on a single variable, self-reported mask use, as an indicator of personal protection measures in the main analyses.

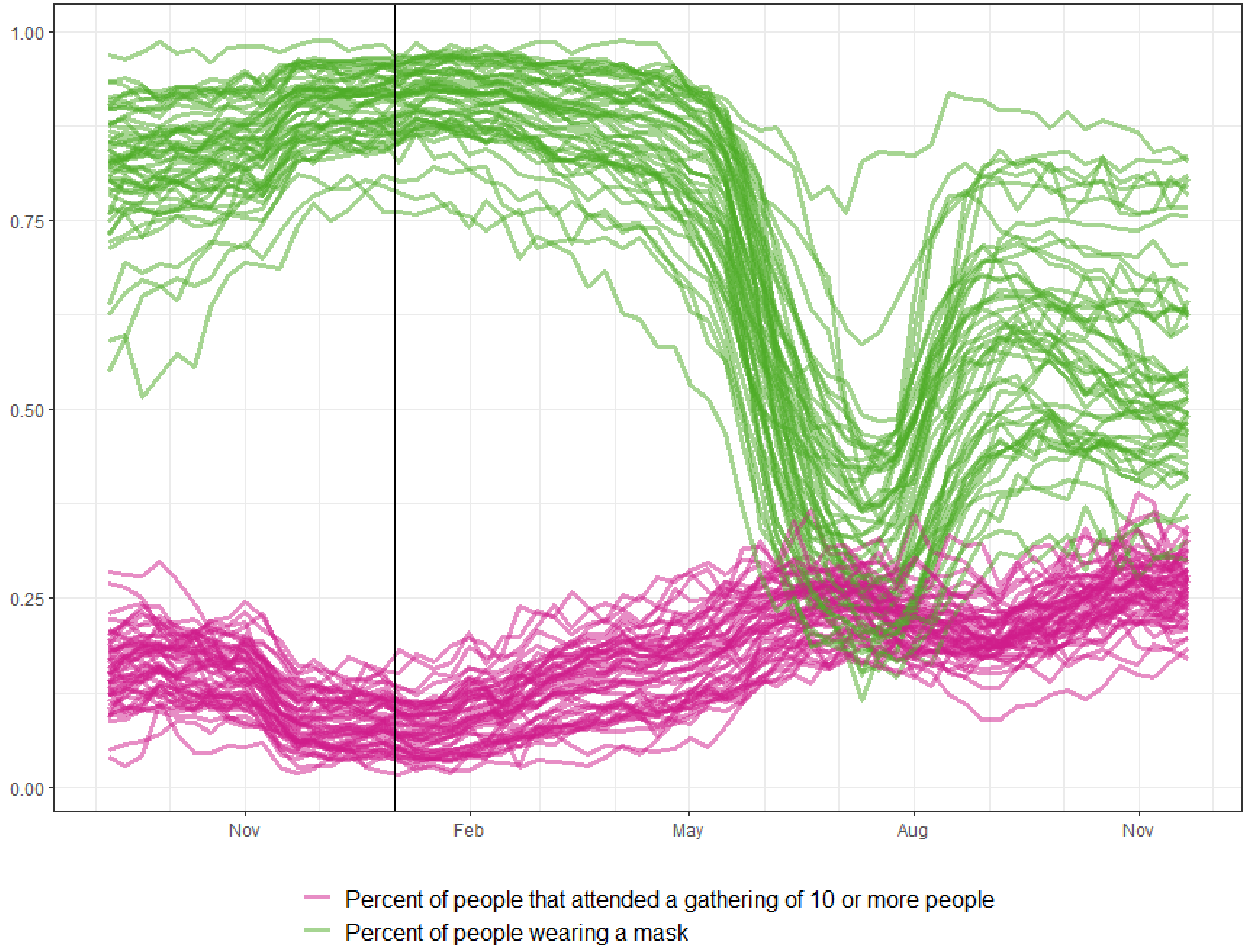

**Fig S3. A. and B.**

We estimated the absolute humidity (AH, grams/meter$^3$) from the daily mean temperature and mean relative humidity (RH) using the Clausius-Clapeyron equation[7,8] (see https://github.com/cdcepi/COVID-19-Mitigation_Rt). **Panel A** shows Pearson correlation coefficients for these three indicators and **panel B** shows the weekly median jurisdiction-level estimates of weather indicators over time, with a solid vertical line representing January 1, 2021. Given the high correlation between AH and temperature, and because we theorize temperature a proxy for both biological mechanisms driving transmission as well as mechanisms driving human behavior, we used only the temperature variable in the regression models. We also excluded RH from the models because it did not add unique information to the results; for example, there was no difference in model fit between models that included RH those that did not, nor was there a difference in the magnitude and direction of any of the covariate posteriors. This is likely due to the null posterior of RH in the model.

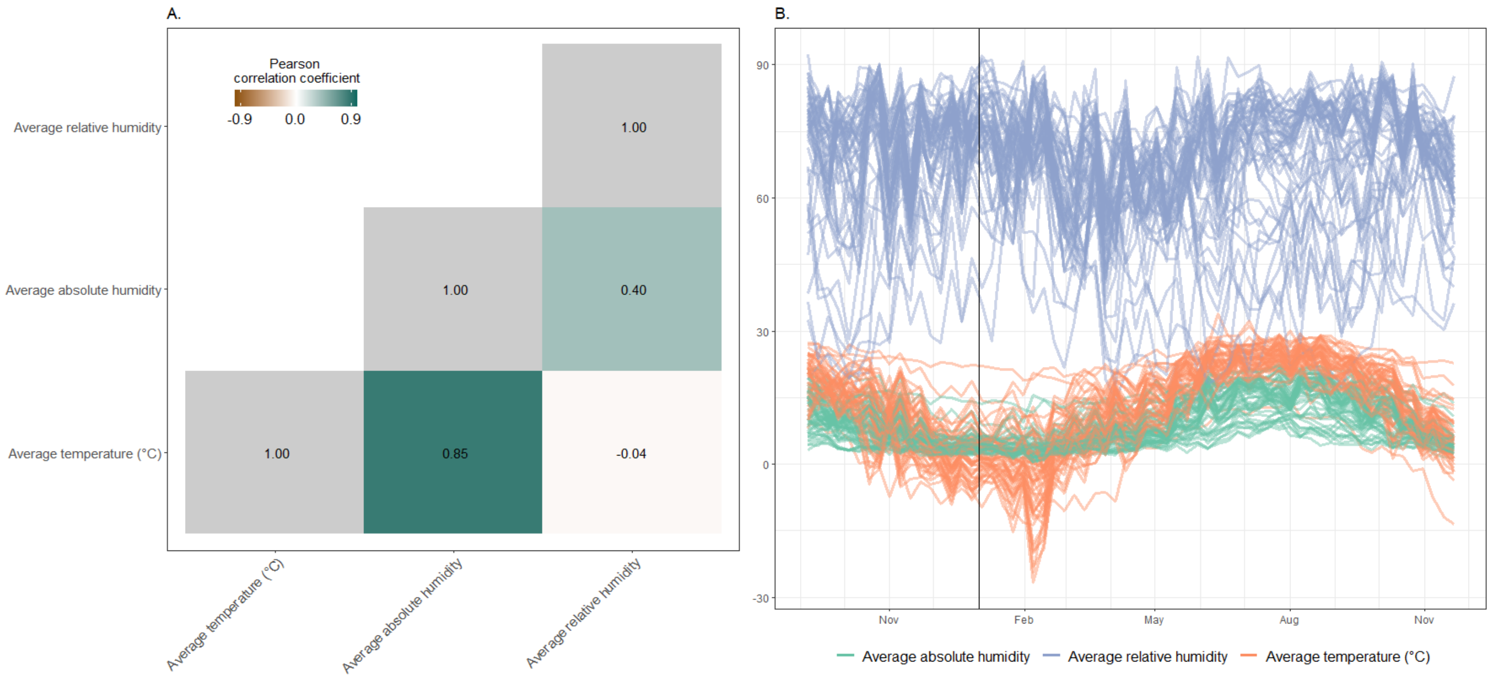


[7] Shi, P. et al. The impact of temperature and absolute humidity on the coronavirus disease 2019 (COVID-19) outbreak - evidence from China. Preprint at *medRxiv* https://doi.org/10.1101/2020.03.22.20038919

[8] Sera, F., Armstrong, B., Abbott, S. *et al.* A cross-sectional analysis of meteorological factors and SARS-CoV-2 transmission in 409 cities across 26 countries. *Nat Commun* **12,** 5968 (2021). https://doi.org/10.1038/s41467-021-25914-8

**Fig S4.**

Pearson correlation coefficients for all Community Covid-19 Vulnerability Index (CCVI) indicators. The CCVI contains seven composite indicators: 1) Racial and Ethnic Diversity (as characterized by Minority Status [defined as an estimate of all persons except white, non-Hispanic individuals] and Language [defined as English language speaking proficiency]), 2) Percentage of Population Working or Living in Environments with High COVID-19 Infection Risk, 3) Socioeconomic Status, 4) Housing type, Transportation, Household Composition and Disability, 5) Population Density, 6) COVID-19 Epidemiological Factors, and 7) Healthcare System Factors. We excluded COVID-19 Epidemiological Factors and Healthcare System Factors from our analysis, as they are likely more closely related to potential disease severity and ability of the health system to respond than drivers of transmission.

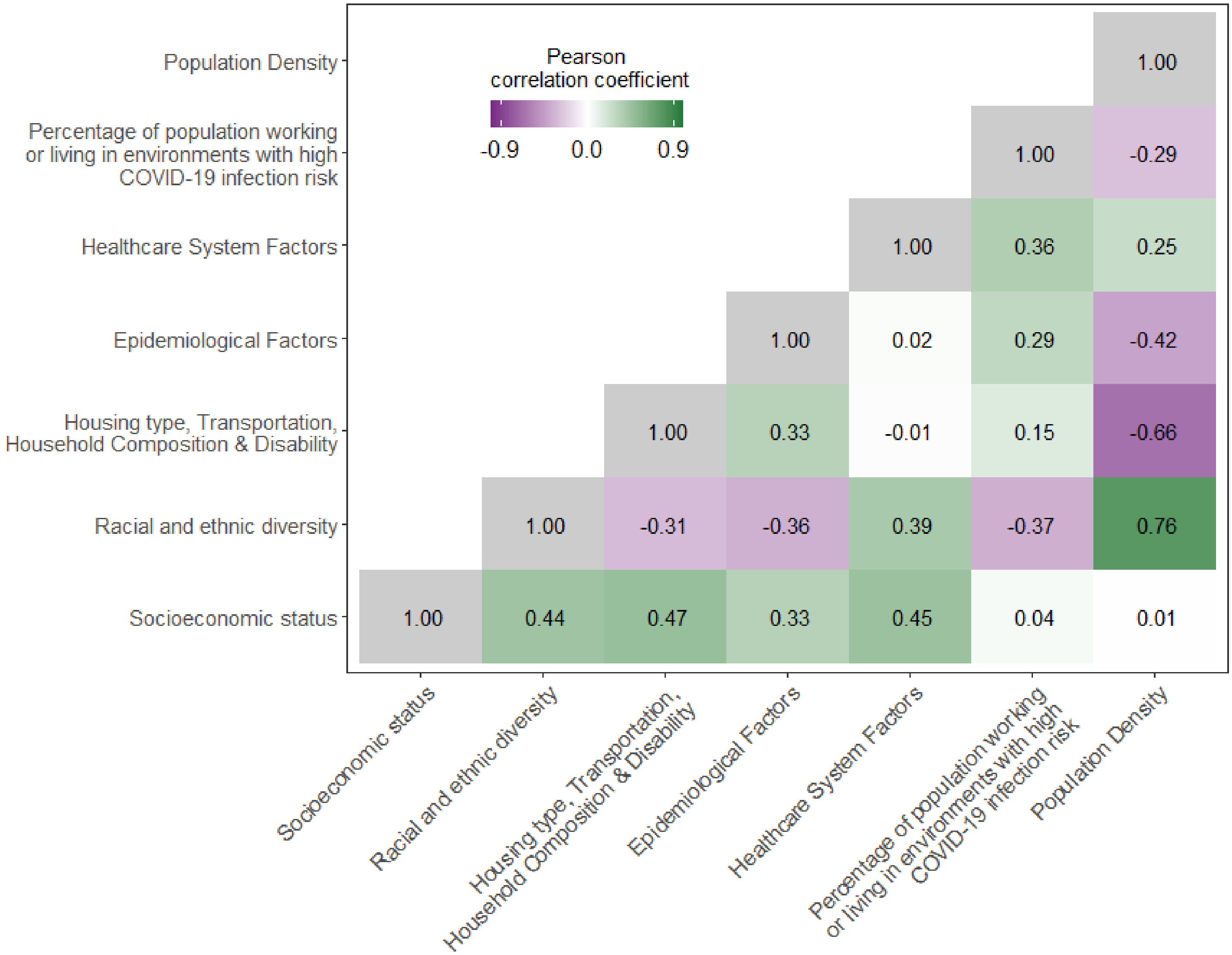

**Fig S5. A. and B.**

**A**. Distribution of Community Covid-19 Vulnerability Index (CCVI) indicators included for each jurisdiction, ordered from highest to lowest average CCVI across the three included indicators. **B**. Pearson correlation coefficient for CCVI indicators and median value for each jurisdiction time-varying variables.

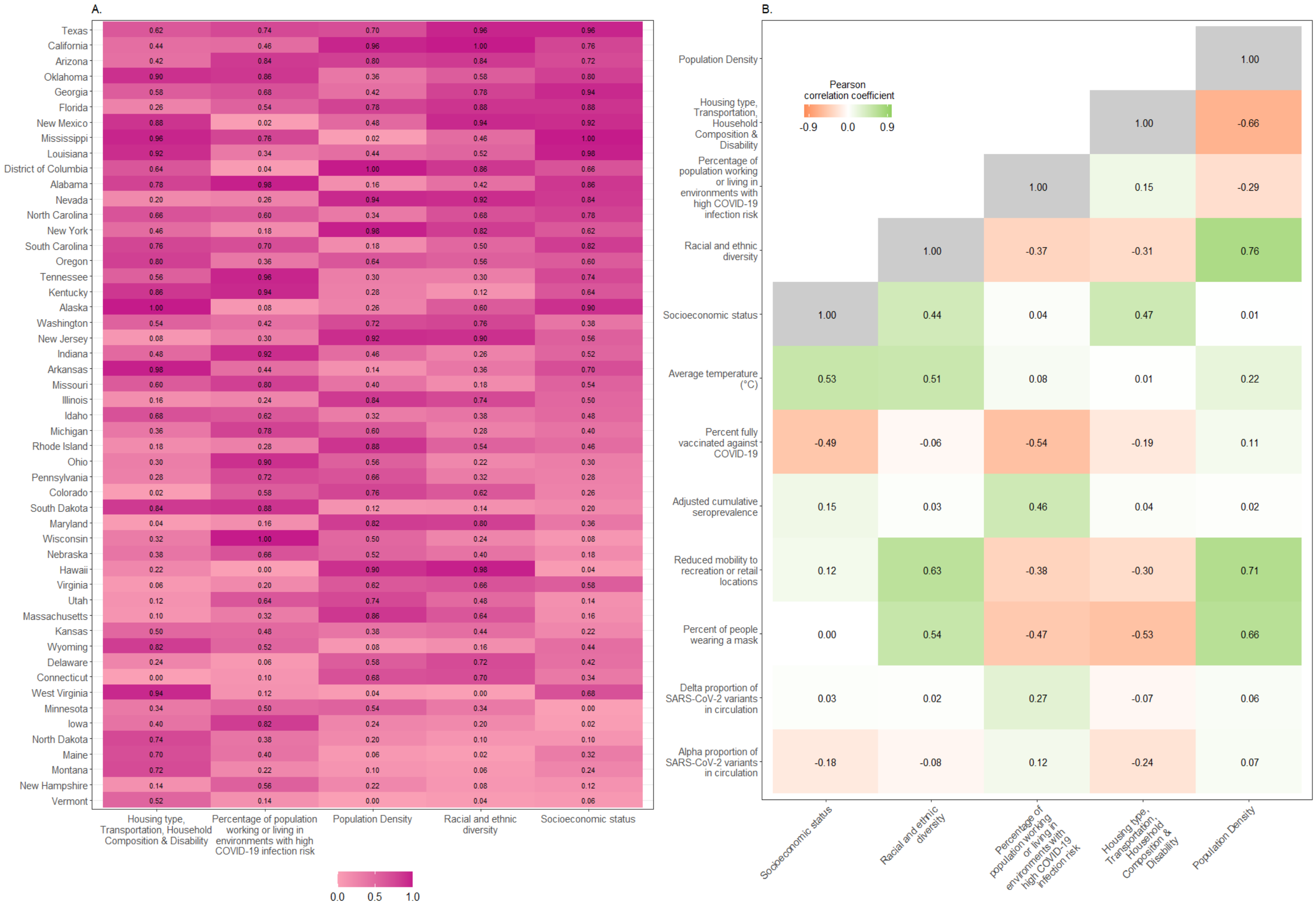

**Fig S6.**

Average relative contribution of observed covariates on fitted COVID-19 time-varying reproduction number (Rt) per jurisdiction. The solid vertical line represents January 1, 2021. The highest values over time (i.e., the top of the pink band) represent $R_t$ estimates including only the effects of local vulnerability and variants. The pink band represents reductions in Rt associated with changing weather. The dark blue band represents reductions in Rt associated with infection induced immunity. The light blue band represents reductions in Rt associated with behavior modification. The light green band represents reductions in Rt associated with policies. The dark green band represents reductions in Rt associated with vaccination. The fitted values from the regression model are represented with the top of the black band.

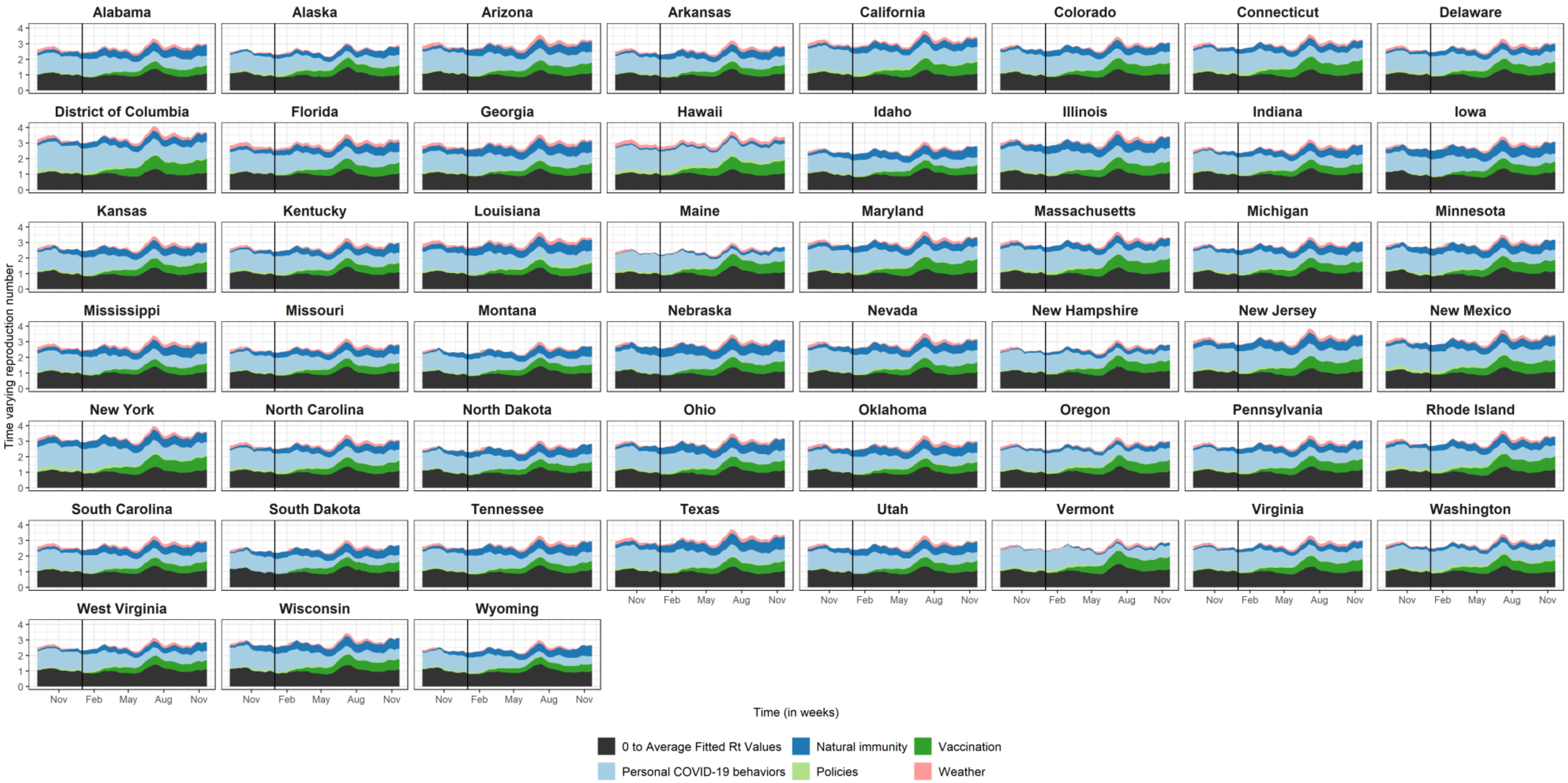

**Fig S7. A. and B.**

**A**. $\hat{R}$ estimates from both OSI models indicated convergence (i.e., values for each covariate were approximate 1.0). Predicted $R_t$ (OSI with specified priors in blue and OSI with naïve priors in red) vs observed values (in grey) are shown for jurisdiction over time, with residuals over time above each $R_t$ plot (OSI model with specified priors in blue and model OSI with naïve priors in red). The solid vertical line represents January 1, 2021. Fit was comparable as there was little difference in predictions or residuals between the models. All posteriors were similar in direction and magnitude. Leave-one-out (LOO) showed non-important differences between the fit of the model with specific priors and the model with naïve priors (-0.4 [2.5 standard error, SE] elpd difference) and Pareto-k^ values for both models suggest limited influence of removing particular observations in LOO.

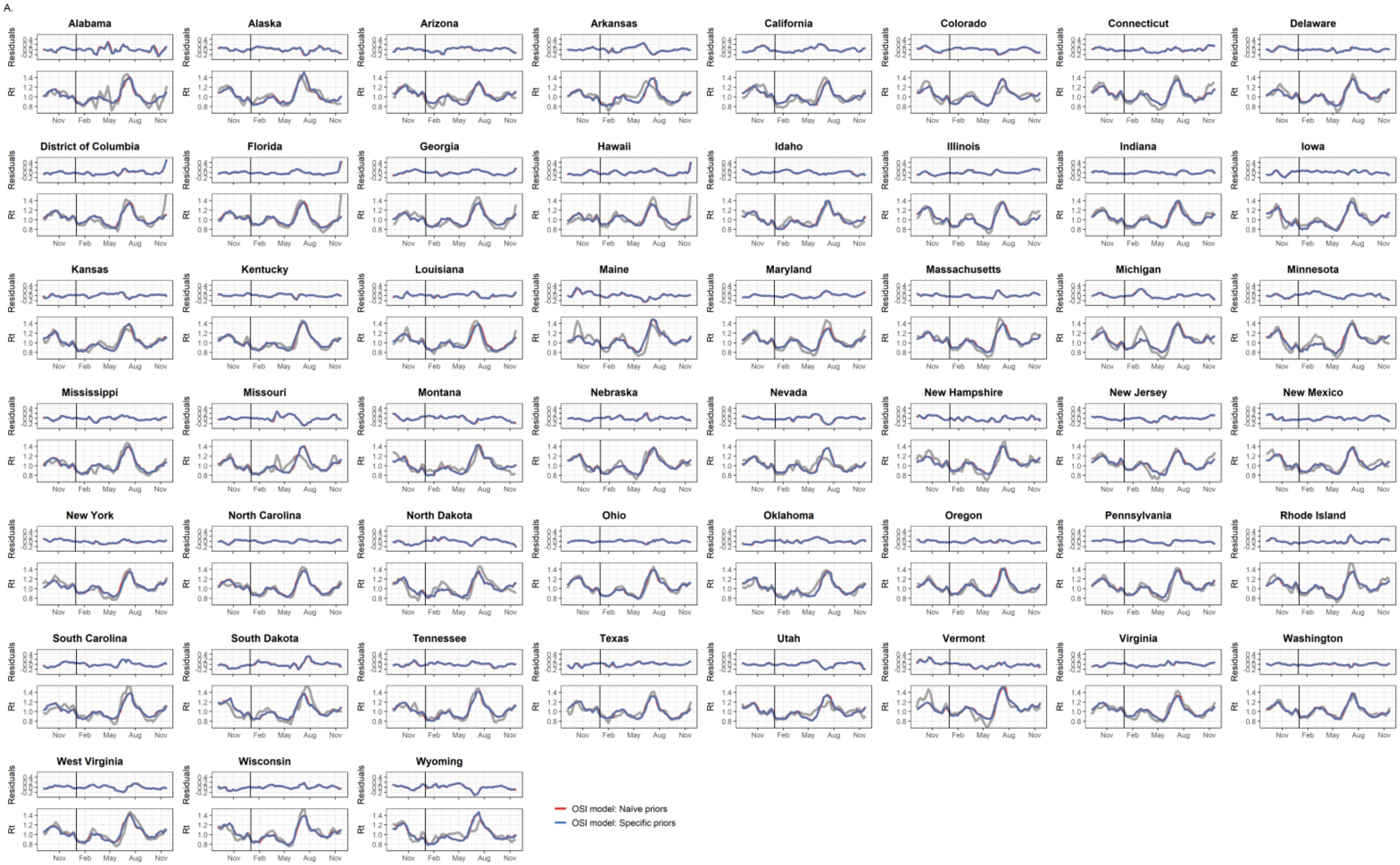

**B.** Both policies models converged as indicated by $\hat{R}$ estimates, and fit was comparable as evident from minor differences in predicted $R_t$ values (model with specified priors in blue and model with naïve priors in red) or residuals. All posteriors were similar in direction and magnitude. Likewise, leave-one-out (LOO) comparison showed nonmeaningful differences between the models (specific prior model had -1.4 [2.7] elpd difference) and acceptable Pareto-k^ values for both models.

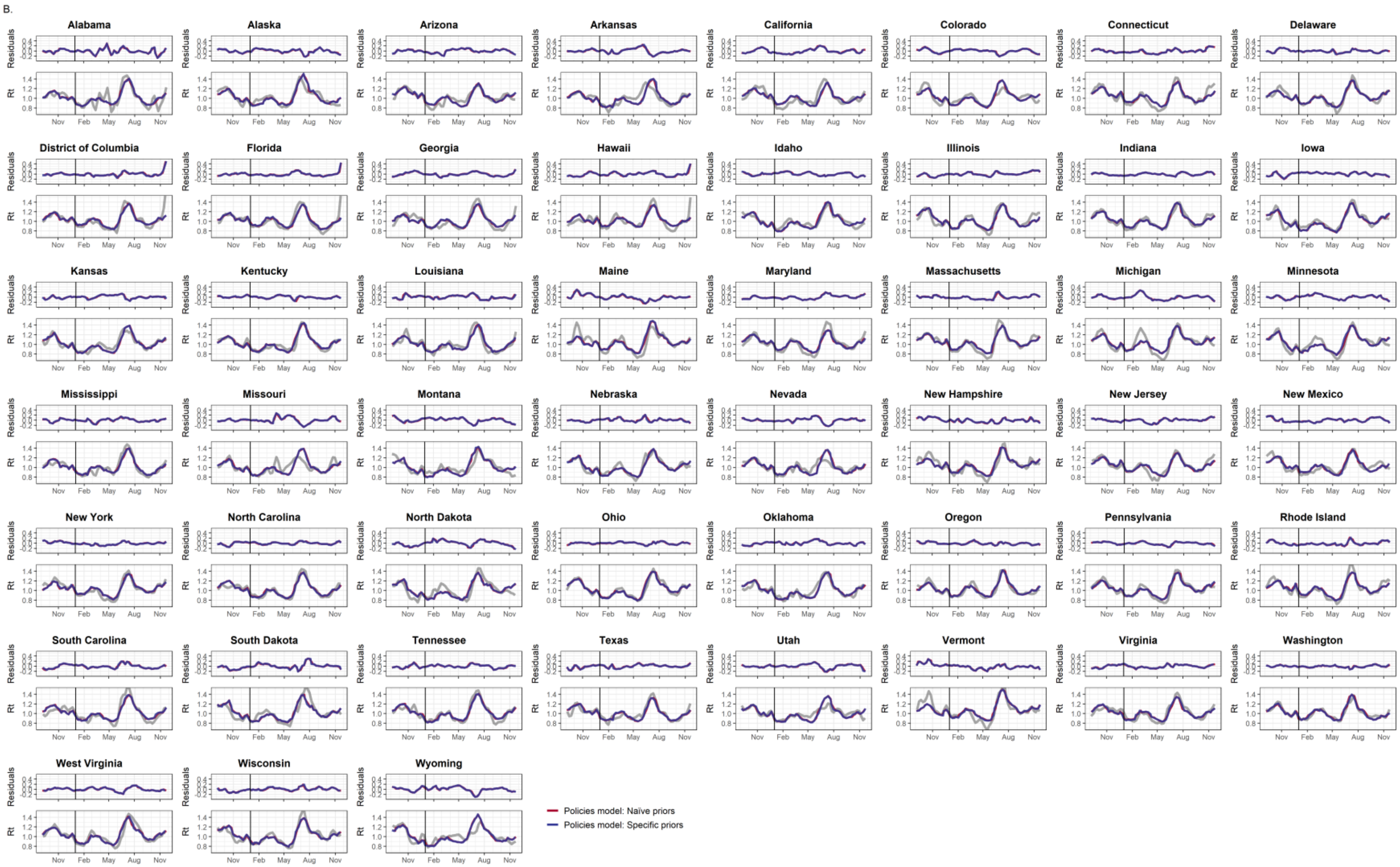

**Fig S8. A. and B.**

**A.** We compared estimated Rt values (in grey dots) to the smooth time from the GAM model (in blue) and the weekly intercepts of the GLM model (in red). The posterior mean is represented by the solid lines and the 95% credible intervals by the bands surrounding it. While the overall time trend was similar for most of the analysis period, the GAM time trend was much lower than the GLM later summer of 2021.

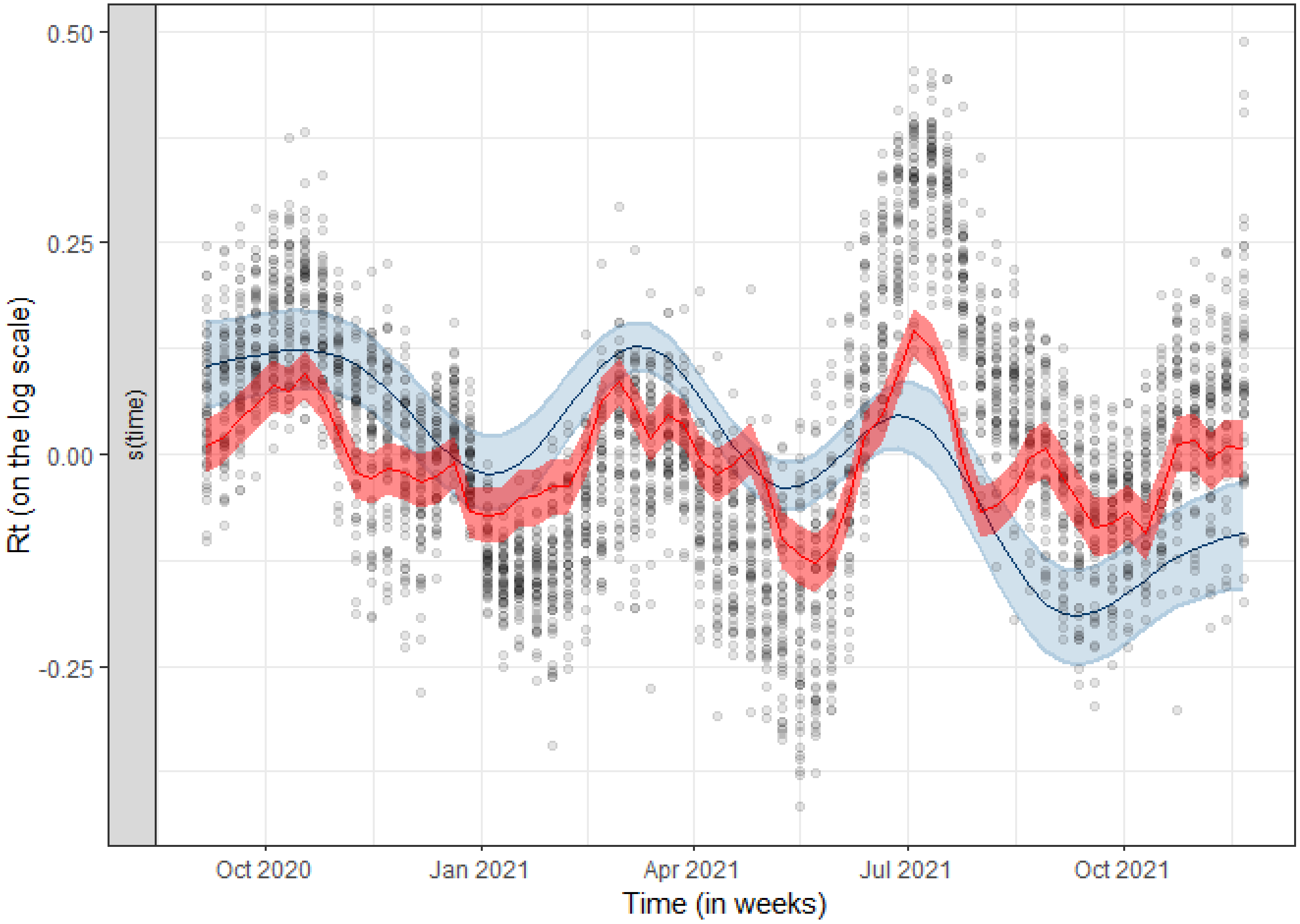

**8B**. $\hat{R}$ estimates from all four models indicated convergence. The figure below shows the percent change in the COVID-19 time-varying reproduction ($R_t$) number from policies, personal COVID-19 behaviors, proportion of key SARS-CoV-2 variants in circulation, weather, immunity to SARS-CoV-2, and variables affecting underlying trends in transmission for each of the four models. The posterior mean is represented by the point and the 95% credible intervals by the bars. While all posteriors were similar in direction between the models, the magnitude of the association between $R_t$ and the percent of people wearing a mask differed, the reduction in national airline travel, and the Delta proportion of SARS-CoV-2 variants in circulation.

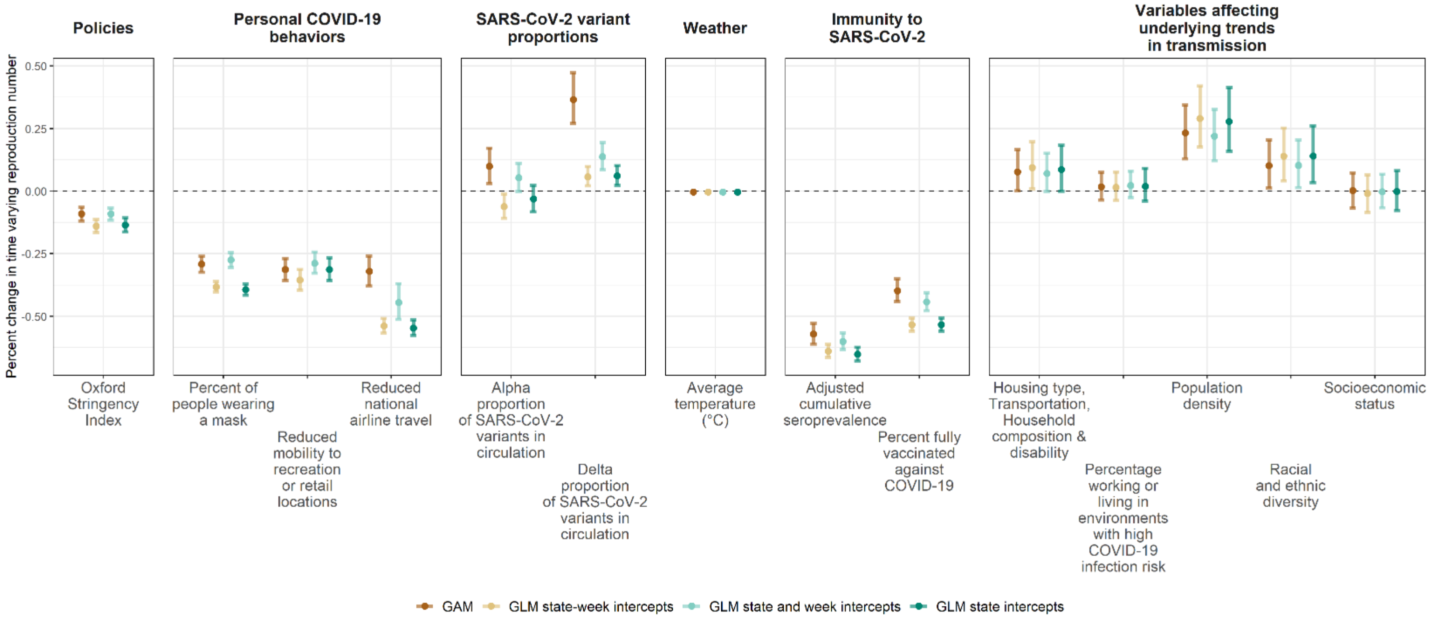

**Fig S9. A., B., and C.**

**A.** Epiforecast $R_t$ estimates (in turquois) and $R_t$ estimates derived from data reported to CDC (in purple) were comparable over time. The 90% uncertainty intervals, as evident in the shaded bans, were wider for summer 2020 peaks in the CDC data. The solid vertical line represents January 1, 2021.

A.

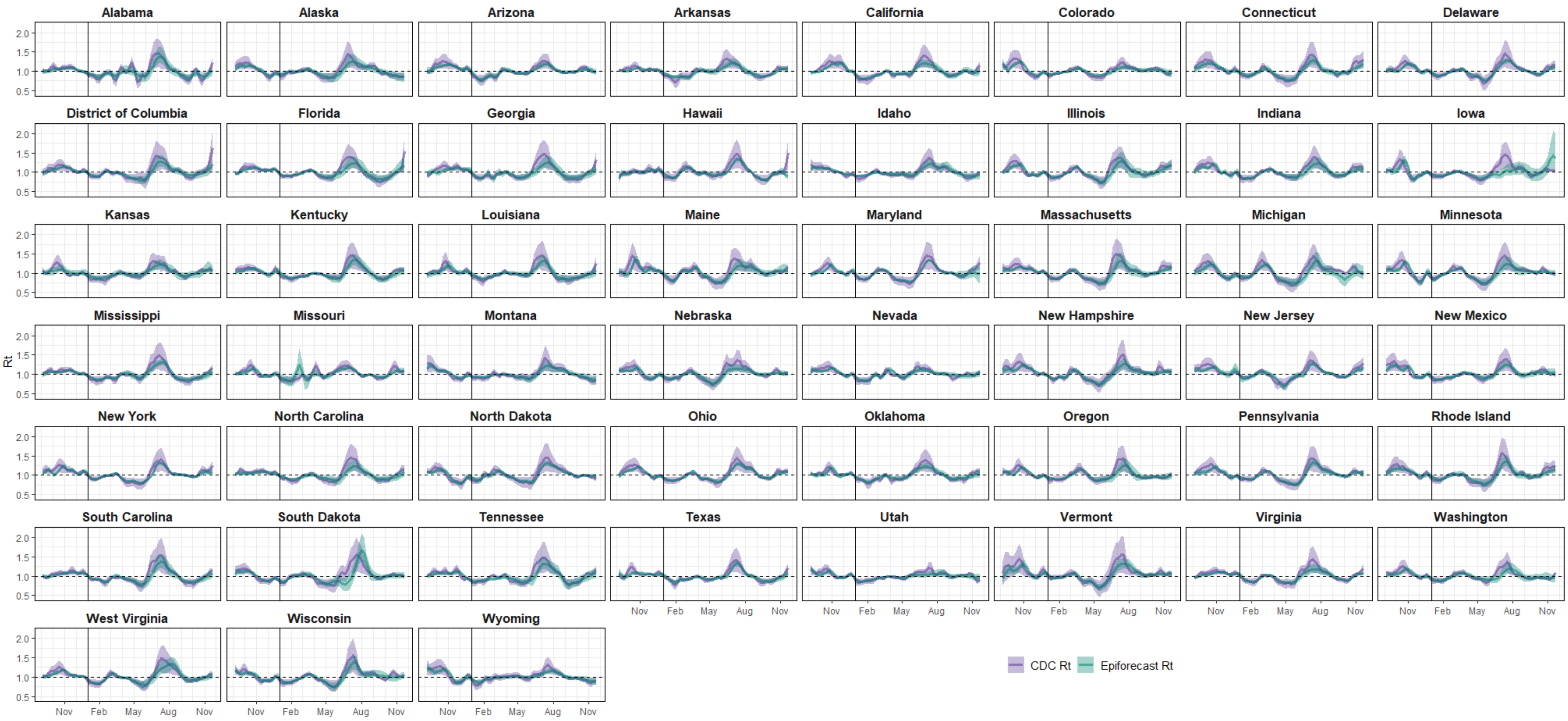

**B.** $\hat{R}$estimates from all four models presented in this sensitivity analysis (i.e., the OSI Model and the Individual Policy Model in the main paper and the models rerun with epiforecast now derived $R_t$) indicated convergence. The figure below shows the percent change in the COVID-19 time-varying reproduction ($R_t$) number from policies, personal COVID-19 behaviors, proportion of key SARS-CoV-2 variants in circulation, weather, immunity to SARS-CoV-2, and variables affecting underlying trends in transmission for each of the four models. The posterior mean is represented by the point and the 95% credible intervals by the bars. All posteriors were similar in magnitude and direction between the two OSI models and the two policy models.

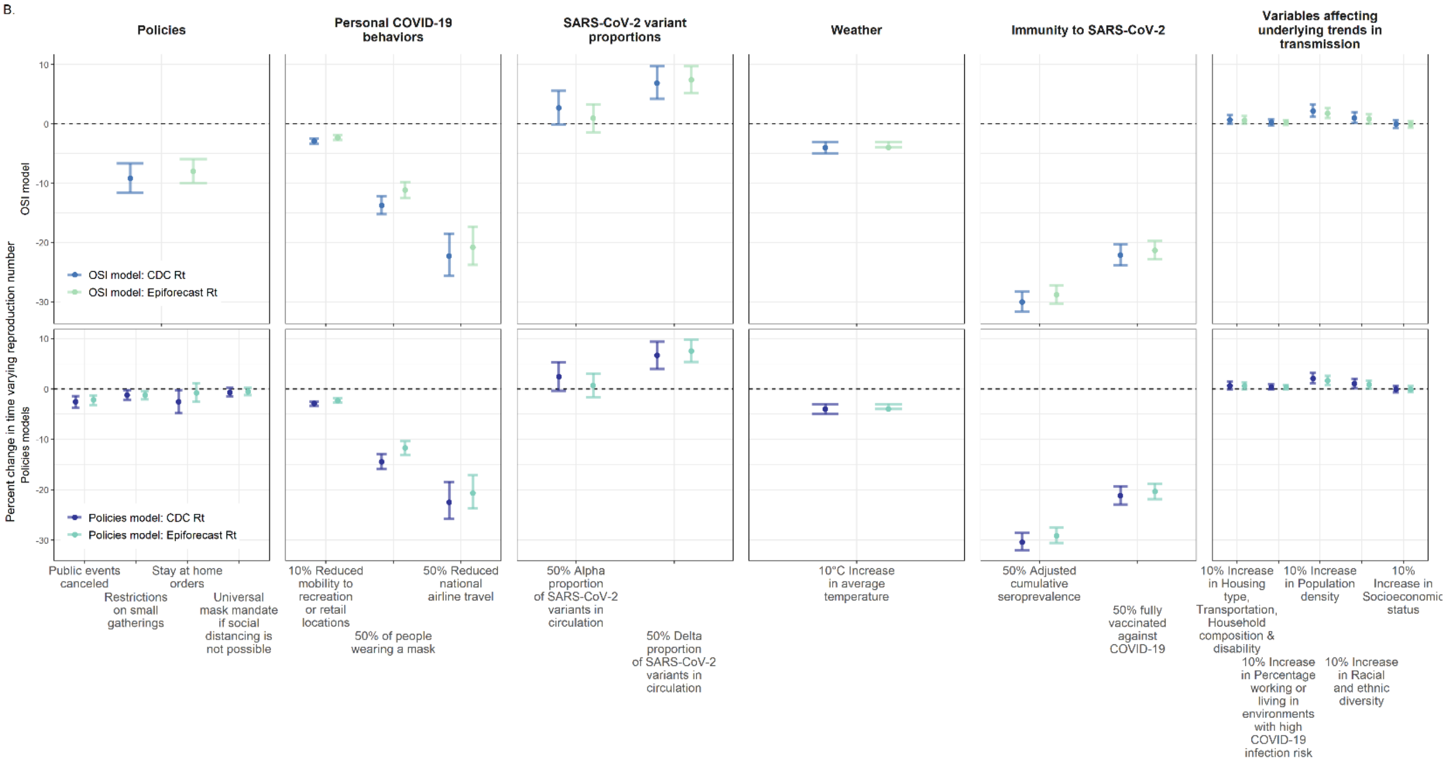

**C**. Per model mean predicted $R_t$ over all jurisdictions vs observed values per jurisdiction (in grey) are shown over time, with residuals per jurisdiction over time above each $R_t$ plot. The solid vertical line represents January 1, 2021. Both models fitted to $R_t$ estimated from CDC data had much higher variance than the models fit to epiforecast $R_t$. Leave-one-out (LOO) estimates between the two sets of models were markedly different. Both the OSI model fit to $R_t$ estimated from CDC data and the policy model fit to $R_t$ estimated from CDC data had much higher LOO estimates (OSI model -644 [75 SE] elpd difference and policy model -643 [74 SE] elpd difference). Pareto-k^ values from the OSI model with CDC data the policy models were within the acceptable range to suggest high accuracy of LOO. Pareto-k^ estimates from the models fit to epiforecast $R_t$ were acceptable, but had 1 observations (< 0.01%) with LOO posteriors that were different from the rest of the sample.

C.

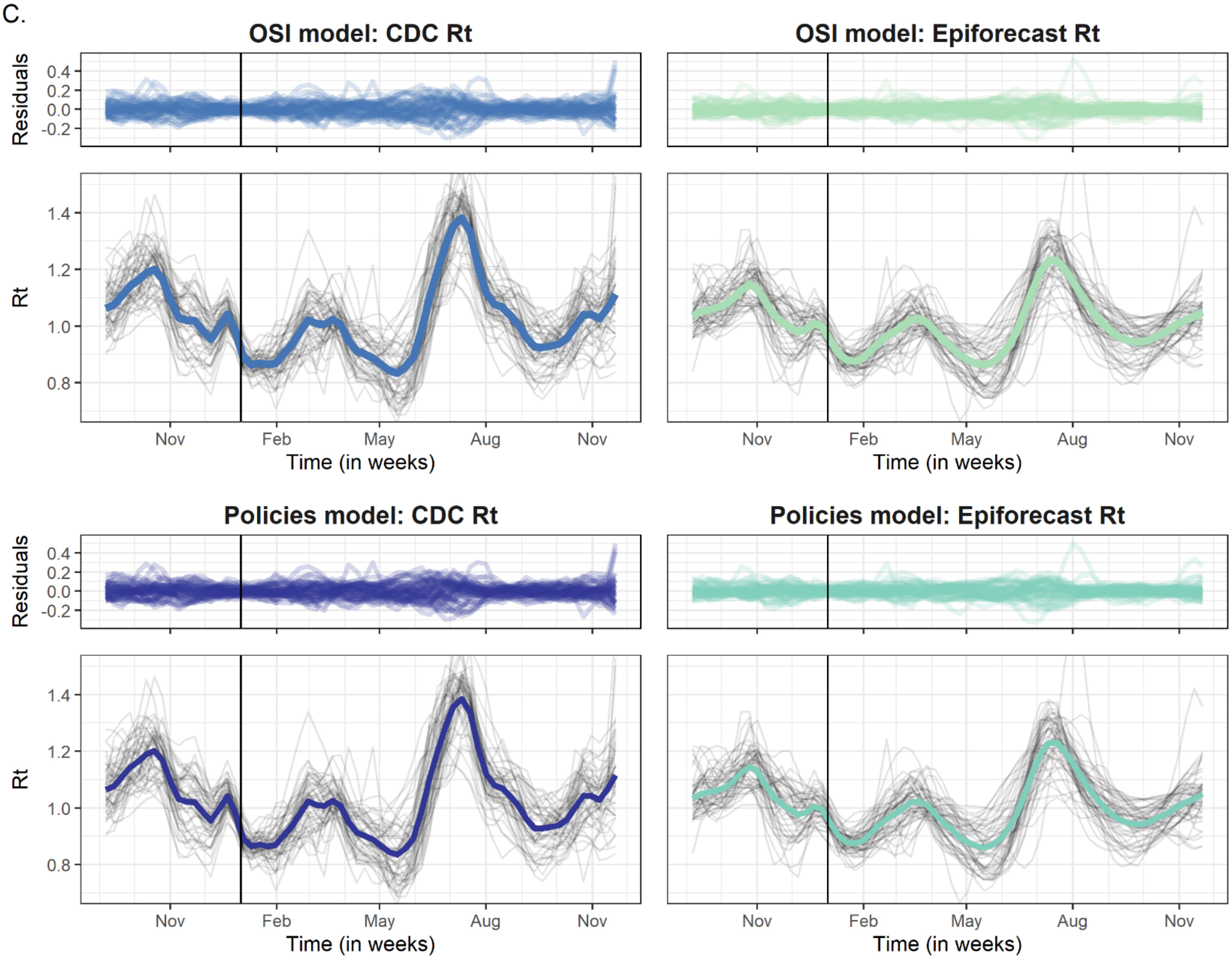

**Fig S10.**

To crudely assess whether the associations between policies and $R_t$ were only driven by personal COVID-19 mitigation behaviors, we re-ran the Individual Policy Model without covariates for mask use or mobility. We then compared the regression posteriors to those from the Individual Policy Model. The figure below shows the percent change in the COVID-19 time-varying reproduction ($R_t$) number from the Individual Policy Model (in orange) and the same model without behavior covariates (in blue). The posterior mean is represented by the point and the 95% credible intervals by the bars. The policy related posteriors variables are not meaningfully different.

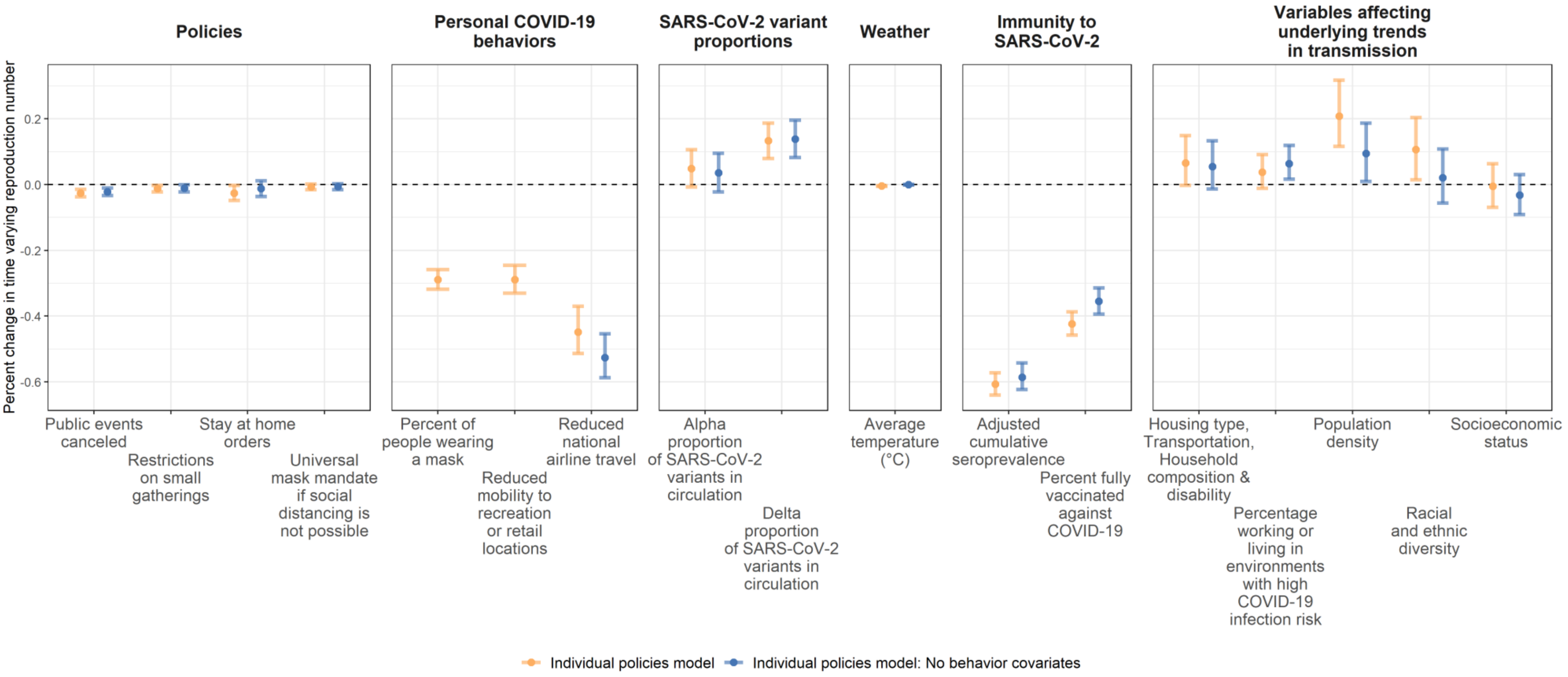

**Table S1.**

**Table S1**. Posteriors estimates and $\hat{R}$ for each regression model.

| Variable category | Variables used in regression models | OSI model | | | | Individual policies model | | | |
|---|---|---|---|---|---|---|---|---|---|
| | | Mean | 2.5% | 97.5% | $\hat{R}$ | Mean | 2.5% | 97.5% | $\hat{R}$ |
| *Policies* | Oxford Stringency Index | -0.096 | -0.123 | -0.069 | 1.001 | - | - | - | - |
| | Public events canceled | - | - | - | - | -0.026 | -0.038 | -0.014 | 1.000 |
| | Stay at home orders | - | - | - | - | -0.012 | -0.023 | -0.002 | 1.000 |
| | Restrictions on small gatherings | - | - | - | - | -0.026 | -0.049 | -0.003 | 1.000 |
| | Universal mask mandate if social distancing is not possible | - | - | - | - | -0.007 | -0.015 | 0.002 | 1.000 |
| *Personal COVID-19 behaviors* | Reduced national airline travel | -0.589 | -0.717 | -0.462 | 1.000 | -0.589 | -0.718 | -0.454 | 1.000 |
| | Reduced mobility to recreation or retail locations | -0.339 | -0.397 | -0.279 | 1.000 | -0.340 | -0.398 | -0.280 | 1.002 |
| | Percent of people wearing a mask | -0.322 | -0.363 | -0.280 | 1.001 | -0.341 | -0.382 | -0.299 | 1.000 |
| *SARS-CoV-2 variant proportions* | Alpha proportion of SARS-CoV-2 variants in circulation | 0.053 | -0.001 | 0.106 | 1.001 | 0.047 | -0.006 | 0.100 | 1.001 |
| | Delta proportion of SARS-CoV-2 variants in circulation | 0.129 | 0.082 | 0.176 | 1.013 | 0.126 | 0.076 | 0.175 | 1.002 |
| *Weather* | Average temperature (°C) | -0.004 | -0.005 | -0.003 | 1.000 | -0.004 | -0.005 | -0.003 | 1.001 |
| *Immunity to SARS-CoV-2* | Adjusted cumulative seroprevalence | -0.919 | -1.002 | -0.835 | 1.002 | -0.935 | -1.019 | -0.849 | 1.001 |
| | Percent fully vaccinated against COVID-19 | -0.584 | -0.650 | -0.522 | 1.001 | -0.550 | -0.612 | -0.490 | 1.001 |

| | | | | | | | | | |
|---|---|---|---|---|---|---|---|---|---|
| | Housing type, Transportation, Household Composition & Disability | 0.068 | -0.001 | 0.141 | 1.001 | 0.064 | -0.005 | 0.141 | 1.001 |
| *Variables affecting the underlying trends in transmission* | Percentage working or living in environments with high COVID-19 infection risk | 0.023 | -0.027 | 0.077 | 1.004 | 0.036 | -0.014 | 0.089 | 1.000 |
| | Population density | 0.199 | 0.116 | 0.283 | 1.004 | 0.191 | 0.114 | 0.279 | 1.002 |
| | Racial and ethnic diversity | 0.098 | 0.014 | 0.186 | 1.005 | 0.098 | 0.016 | 0.181 | 1.002 |
| | Socioeconomic status | -0.002 | -0.068 | 0.065 | 1.002 | -0.001 | -0.066 | 0.065 | 1.004 |

**Table S2.**

**Table S2**. Elpd from 10-fold CV for each of the four models

| Model | 10-fold elpd (standard error) |
|---|---|
| GLM: state random intercept only | 3092 (52) |
| GLM: random intercepts per state-week | 5992 (14)* |
| GLM: random intercepts per state and per week | 3803 (61) |
| GAM: random intercepts per state | 2667 (67) |

**Only model with Pareto-k^ estimates indicating LOO posteriors that were importantly different from the rest of the sample, i.e. only 1% of observations feel into an acceptable Pareto-k^ range.*
